# Reference calibration, autozygosity and blood-derived variant interpretation in older adults

**DOI:** 10.64898/2026.10.28.26366687

**Authors:** Myriam El-Haddad, Nagham N. Hendi, Ghada El-Hajj Fuleihan, Georges Nemer, Nehme El-Hachem

## Abstract

Population exomes contain signals of demographic history, inherited disease alleles and, when derived from blood, possible acquired variation. Distinguishing these signals is important in underrepresented populations, where reference absence can reflect both sampling and local frequency differences. To the best of our knowledge, this study represents the first Levantine old-age cohort characterized by whole exomes. We reanalyzed blood-derived exomes from 199 Lebanese trial participants aged 65–91 years, including 187 unrelated participants for frequency and recessive analyses. Twenty-nine participants (14.6%) reached the autozygosity expected for offspring of first cousins. Although 13.8% of observed autosomal alleles were absent at callable gnomAD Middle Eastern reference sites, size matching gave a reference-to-cohort single-nucleotide variant absence ratio of 0.98. Excess absence was concentrated among recurrent alleles; 360 were absent from the full reference, none classified as pathogenic or likely pathogenic in ClinVar. Separately, regional disease alleles showed nominal frequency differences, including MEFV p.Val726Ala (0.051 versus 0.012). Qualifying recessive alleles occurred in 55.1% of unrelated participants, decreasing to 45.5% after severity-related exclusions. Conditional qualifying biallelic genotype estimates increased from 2.0 to 27.7 per 1,000 offspring between unrelated-parent and first-cousin inbreeding coefficients. Twelve participants had clonal-driver candidates, but only five met both binomial and empirical allele-fraction criteria for the probable tier; a TP53 frameshift remained unresolved. Eight participants (4.0%) had unconfirmed, condition-matched candidate secondary findings. These observations distinguish reference sampling from allele-specific frequency differences and genotype expectations from clinical outcomes. They support complementary checks on population exome interpretation, while leaving generalizability, variant origin and clinical reportability to independent evaluation.

**Author Summary:** We studied genetic variation in 199 older adults from Lebanon to understand how population differences and aging can complicate the interpretation of genetic tests. A change missing from a reference database might be locally common, but it might also be missing simply because the database sampled too few people. We compared equally sized samples and found that the overall absence of observed changes was similar in the regional reference and another part of our study group. Individual disease-associated changes could nevertheless have different local frequencies. We also examined stretches of DNA inherited from shared ancestors and estimated how parental relatedness could change the chance of pairing disease-associated copies. These estimates describe genetic combinations, not predicted numbers of affected children. Finally, because blood cells can acquire changes during aging, we compared candidate changes with the sequencing patterns of other variants in the same data. Some findings remained uncertain, and none was clinically confirmed. Our results show why reference sampling, shared ancestry and possible acquired changes need separate consideration before drawing conclusions about inherited risk.

## Introduction

Genetic variation is shaped by population history, but the variation observed in a sequencing study also depends on who was sampled, which genomic regions were assayed and how variants were detected. These influences become difficult to separate when a cohort comes from an underrepresented population. Arab populations remain sparsely represented in genomic studies [1], and the gnomAD v4.1 Middle Eastern group combines approximately 3,000 individuals without a separate Levantine reference [2]. A regional cohort can therefore reveal useful allele observations while also posing a statistical question: which apparent differences reflect local frequency structure, and which are expected consequences of comparing finite samples?

Reference absence is one example. An allele absent from an adequately assayed reference site might be geographically concentrated, but it might instead be rare enough to escape another sample from the same population. These possibilities cannot be distinguished from the raw proportion of reference-absent alleles alone. A comparison between equally sized samples supplies an internal benchmark: does the regional reference miss observed cohort alleles more often than another subset of the cohort does? This question concerns the representation of observed exome alleles, not whether the reference contains all genomic diversity. The distinction matters because allele-frequency evidence also informs disease-variant interpretation [3–4].

Autozygosity introduces a related but different consequence of population history. Genome segments inherited from a shared ancestor can increase the probability that two copies of a recessive allele meet in an offspring, without necessarily increasing the proportion of individuals carrying any qualifying allele. Consanguinity and genome-wide inbreeding have been documented in Lebanon [5–6], as have founder haplotypes for regional disease alleles [7]. An explicit inbreeding model can connect observed alleles to expected biallelic genotypes, but that expectation is not equivalent to disease incidence. Variant severity, penetrance, phase and alleles not observed in a small cohort remain relevant.

Age and tissue source add another layer. Expanded hematopoietic clones introduce acquired driver variants into blood-derived sequence data [8–9]. Allelic imbalance may suggest a clonal origin, but technical effects also displace constitutional heterozygotes from a fraction of 0.5. Blood-detected *TP53* variants illustrate the resulting ambiguity between acquired and inherited findings [10]. Comparing candidates with an empirical allele-fraction background can contextualize imbalance, although blood alone cannot resolve variant origin.

Here we reanalyzed exomes previously generated from older Lebanese participants in a vitamin D trial [11–12]. We examined autozygosity, size-matched reference absence, regional disease alleles, conditional recessive genotype burden and candidate clonal and inherited-risk findings. Size matching substantially changed the interpretation of overall reference absence, whereas allele-specific frequency differences and uncertain blood-derived findings remained. The study uses these observations to distinguish sampling, local allele frequencies, shared ancestry and possible acquired variation within one selected cohort; it does not establish a population-representative resource or validate a clinical decision rule.

## Results

### Cohort structure and autozygosity

After joint calling against GRCh38 and genotype filtering for read support and excess heterozygosity (Materials and Methods), call rates ranged from 92.9% to 98.2% (median 97.2%) and mean depth from 25.9× to 38.0× (Supplementary Table 17). The cohort lay near European and Middle Eastern references on the first principal components (Fig. 1b,c), and all 199 participants were closest to the Middle Eastern centroid when five or more components were included (Supplementary Fig. 2). Removing one member of each of 15 related pairs left 187 unrelated participants for frequency analyses (Table 1).

**Fig 1.**
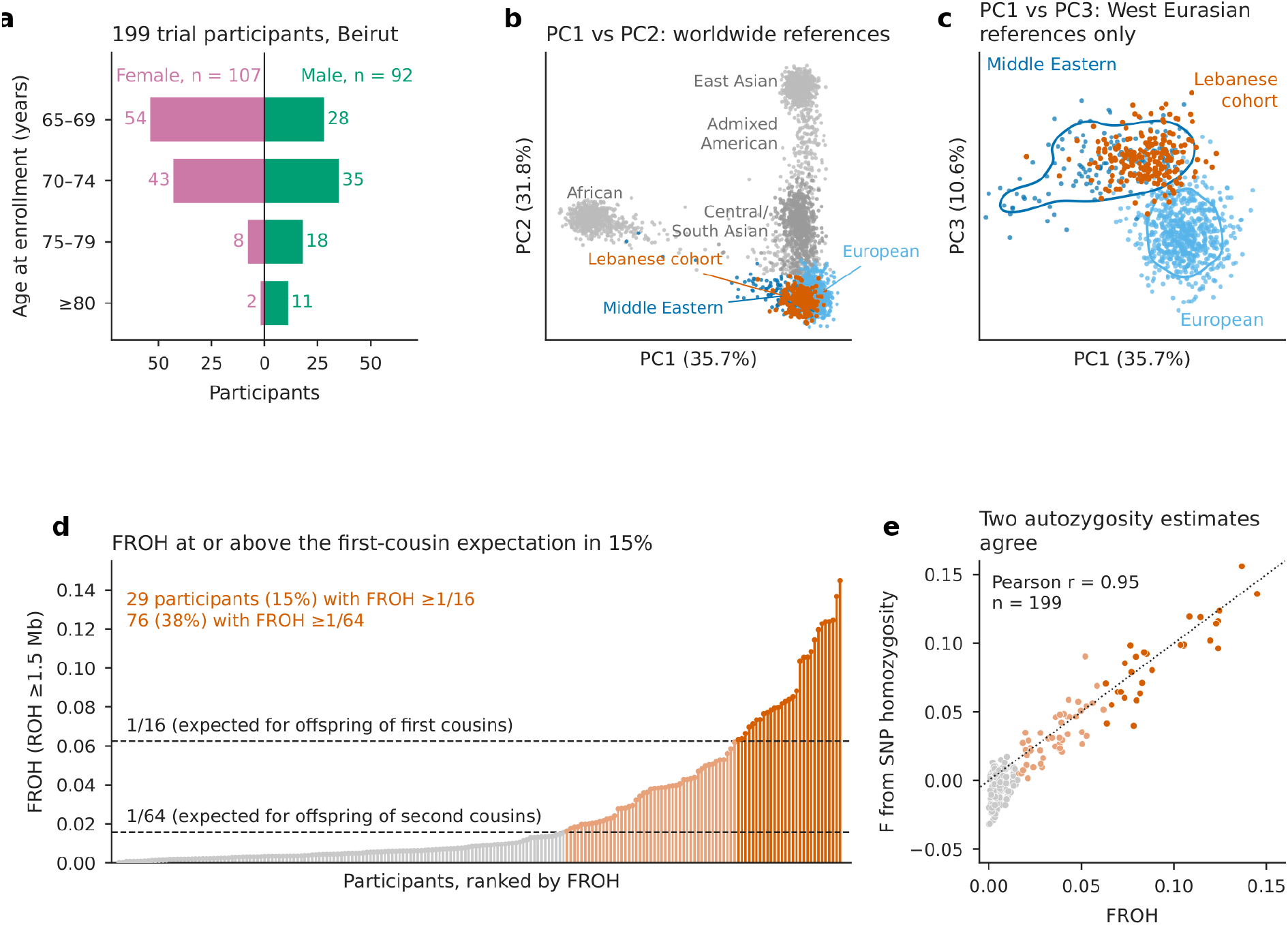
Cohort, ancestry and autozygosity. **a**, Age groups and recorded sex. **b**, Joint principal-component analysis of 199 participants (vermilion) and 3,280 HGDP/1000 Genomes references; European and Middle Eastern groups are highlighted. **c**, West Eurasian projection with contours enclosing the densest 75% of each reference group; this is not an enlargement of **b**. Axis percentages refer to variance among the first ten components. **d**, Ranked autosomal fraction in runs of homozygosity ≥1.5 Mb (F_ROH_); dashed lines mark first-cousin (1/16) and second-cousin (1/64) expectations. **e**, SNP-based inbreeding coefficient versus F_ROH_; dotted line, identity.

**Table 1.** Cohort and principal findings.

| Measure | Value | Note |
|---|---|---|
| Participants | 199 | Vitamin D trial participants with exomes; Beirut, 2011–2013 |
| Age, years | 71.2 ± 4.8 | Mean ± s.d.; range 65–91 |
| Recorded sex | 107 female; 92 male | 3 discordant with genetic sex |
| Relatedness | 15 related pairs | 187 unrelated for frequency, calibration and recessive analyses; 199 for cancer, clonal, SF and *MEFV* proportions |
| F<sub>ROH</sub> ≥ 1/16 | 29 (14.6%) | 23–29 across methods; mean F<sub>ROH</sub> 2.5% |
| Callable absence from full MID | 13.8% | 187 unrelated; autosomal; unmatched; 1.5% expected under beta-binomial model |
| Size-matched absence, MID versus cohort half | Ratio 0.98 (SNVs) | 0.97–0.99 across ten splits; excess only for alleles seen ≥6 times (ratio 2.1–2.8) |
| Frequency contrasts versus MID | 474 higher; 287 lower | 93,724 coding/splice variants; *P* < 5.3 × 10<sup>−7</sup> |
| Classic *MEFV* exon 10 carriers | 29 (14.6%) | p.Val726Ala allele frequency 0.051 versus 0.012 in MID |
| Carriers of severe-recessive alleles | 103 of 187 (55.1%) | 85 (45.5%) excluding variable-severity alleles |
| Modeled first-cousin excess, per 1,000 offspring | 25.8 (21.7–29.9) | Conditional model output; 18.7 after exclusions |
| Clonal hematopoiesis, probable / any candidate | 5 (2.5%) / 12 (6.0%) | *TP53* frameshift possible; origin unresolved |
| Candidate findings in established dominant cancer genes | 4 (2.0%) | *PALB2*, *PMS2*, *SDHB*, *ATM*; unconfirmed |
| Candidate ACMG SF v3.3 findings | 8 (4.0%) | 95% CI 2.1–7.7%; condition-matched; none confirmed |

Germline values use genotypes filtered for read support and excess heterozygosity; clonal hematopoiesis used a separate somatic branch without an allele-fraction floor. F<sub>ROH</sub>, fraction of the autosomes in runs of homozygosity; MID, gnomAD v4.1 Middle Eastern group; SF, secondary finding.

Mean F_ROH_, the fraction of the autosomes in runs of homozygosity of at least 1.5 Mb, was 2.5% (median 0.9%, maximum 14.5%), in agreement with the 2.3% mean inbreeding estimated from SNP arrays in Lebanese communities [6]. Twenty-nine participants (14.6%) reached the value expected for offspring of first cousins (F_ROH_ ≥ 1/16) and 76 (38.2%) the second-cousin value (≥1/64; Fig. 1d). Single-nucleotide polymorphism (SNP)-based *F* was strongly correlated with F_ROH_ (*r* = 0.95; Fig. 1e), and 23 to 29 participants reached the first-cousin expectation across alternative methods addressing exome-marker gaps (Supplementary Table 13). F_ROH_ was not associated with age (Spearman *ρ* = −0.06, *P* = 0.43).

### Size matching calibrates observed reference absence

Of 575,436 filtered alleles in all 199 participants, 78,039 (14.1% of those with a Middle Eastern allele number) were absent from a callable site in the gnomAD Middle Eastern group (MID; Supplementary Table 20). Among the 187 unrelated participants, 13.8% of autosomal alleles were absent, against 1.5% expected under a beta-binomial sampling model with a Jeffreys prior. Taken at face value, this ninefold excess would suggest extensive population-specific variation.

The parametric model, however, was itself poorly calibrated. Applied within the cohort, it predicted that 35% of singletons in one half would be absent from the other half, whereas 72% were (Supplementary Table 21). We therefore replaced it with an empirical, size-matched comparison. We split the unrelated participants at random into halves of 94 and 93 people ten times and recorded, for each allele seen in the first half, whether it was absent from the 186 allele copies of the second half and the probability that it would be absent from 186 allele copies drawn at random from MID. If the reference represented the cohort poorly, the MID subsample should miss more cohort alleles than the cohort replicate does.

It did not. At autosomal single-nucleotide variants (SNVs), the MID subsample lacked cohort alleles at the same rate as the other half of the cohort (ratio 0.98; range 0.97–0.99 across splits; Fig. 2a). The ratio remained between 0.98 and 1.17 for alleles seen up to five times at sites callable in every member of the comparison half (Supplementary Fig. 14 and Supplementary Table 21c). An excess was confined to recurrent alleles: among alleles seen at least six times, 0.33% of SNVs at well-called sites were absent from the MID subsample against 0.13% from the cohort replicate (ratio 2.1–2.8 across splits). Of these recurrent alleles, 360 were absent from the full MID group, none of them classified as pathogenic or likely pathogenic (P/LP) in ClinVar. In this size-matched analysis of observed exome alleles, overall SNV absence was similar between MID and the cohort comparison, with excess absence concentrated among recurrent alleles. This does not assess unobserved genomic variation.

**Fig 2.**
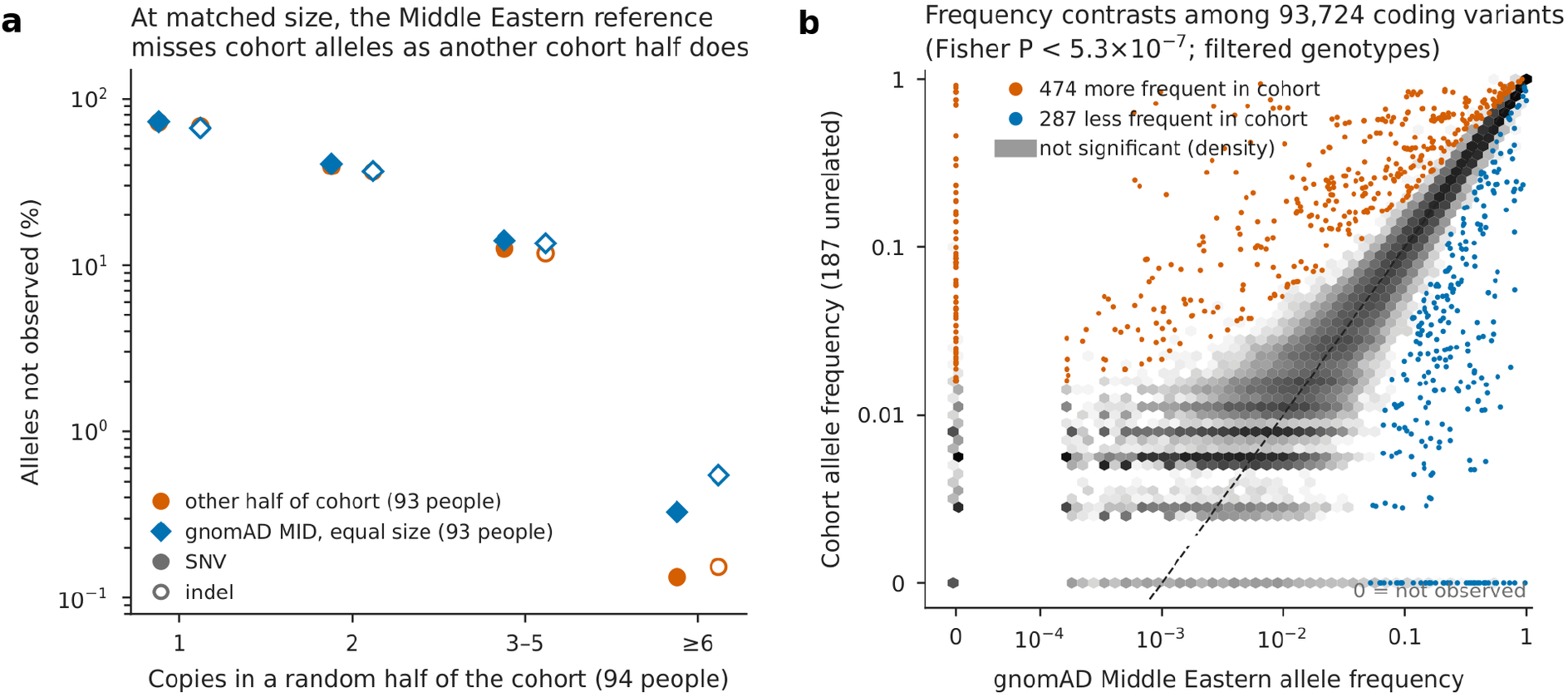
Size-matched reference absence and allele-frequency comparisons. **a**, Alleles observed in 94 discovery participants but absent from 93 comparison participants (186 allele copies; vermilion circles), or their hypergeometric absence probability in 186 gnomAD Middle Eastern (MID) copies (blue diamonds), by discovery count. Filled symbols indicate SNVs; open symbols indicate indels. Points are means of ten overlapping splits, not independent replications. Sites were callable at depth ≥10 in ≥189/199 participants; the fully callable comparison is in Supplementary Fig. 14. **b**, Frequencies in 187 unrelated participants versus MID for 93,724 filtered coding/splice variants. Higher (vermilion) and lower (blue) frequencies meet P <0.05/93,724; zero frequencies occupy positions marked 0.

### Allele-specific frequency differences

Allelic tests of 93,724 coding and splice variants identified 474 that were more frequent and 287 that were less frequent in the unrelated participants than in MID (*P* < 5.3 × 10^−7^; Fig. 2b). Most apparent differences in unfiltered data arose at poorly supported sites (Supplementary Figs. 3–5). For example, three P/LP-annotated alleles in *CNN2* and *VWF* were heterozygous in 74% to 80% of participants with no homozygotes, a pattern compatible with mismapped paralogous reads [13], and were removed by the excess-heterozygosity filter (Supplementary Fig. 6).

Among pre-specified Eastern Mediterranean disease alleles (Fig. 3a), *MEFV* p.Val726Ala had an allele frequency of 0.051 (19 of 374 chromosomes), four times the MID value (0.012; *P* = 1.3 × 10^−6^), and p.Met680Ile was about seven times more frequent (0.013 versus 0.0020; *P* = 0.0022). Twenty-nine participants (14.6%) carried at least one of six classic *MEFV* exon 10 variants, and two unrelated participants carried two (phase undetermined). *SMPD1* p.Leu163Pro, a Niemann–Pick type A/B allele, occurred on 3 of 374 chromosomes compared with 2 of 5,958 in MID (*P* = 0.0019), a more than 20-fold difference. Alleles for cblC methylmalonic aciduria (*MMACHC* p.Val183ThrfsTer5; *P* = 7.5 × 10^−4^), hyperphenylalaninemia (*PAH* p.Ala300Ser; *P* = 0.0038) and hereditary fructose intolerance (*ALDOB* p.Ala338Val; *P* = 0.0088) were also nominally enriched, whereas the *G6PD* Mediterranean allele was less frequent (1.4% versus 4.2% of X chromosomes; *P* = 0.013). These *P* values are nominal for a pre-specified set and are not corrected for multiplicity; the *MEFV* p.Val726Ala comparison did not meet the genome-wide threshold above.

**Fig 3.**
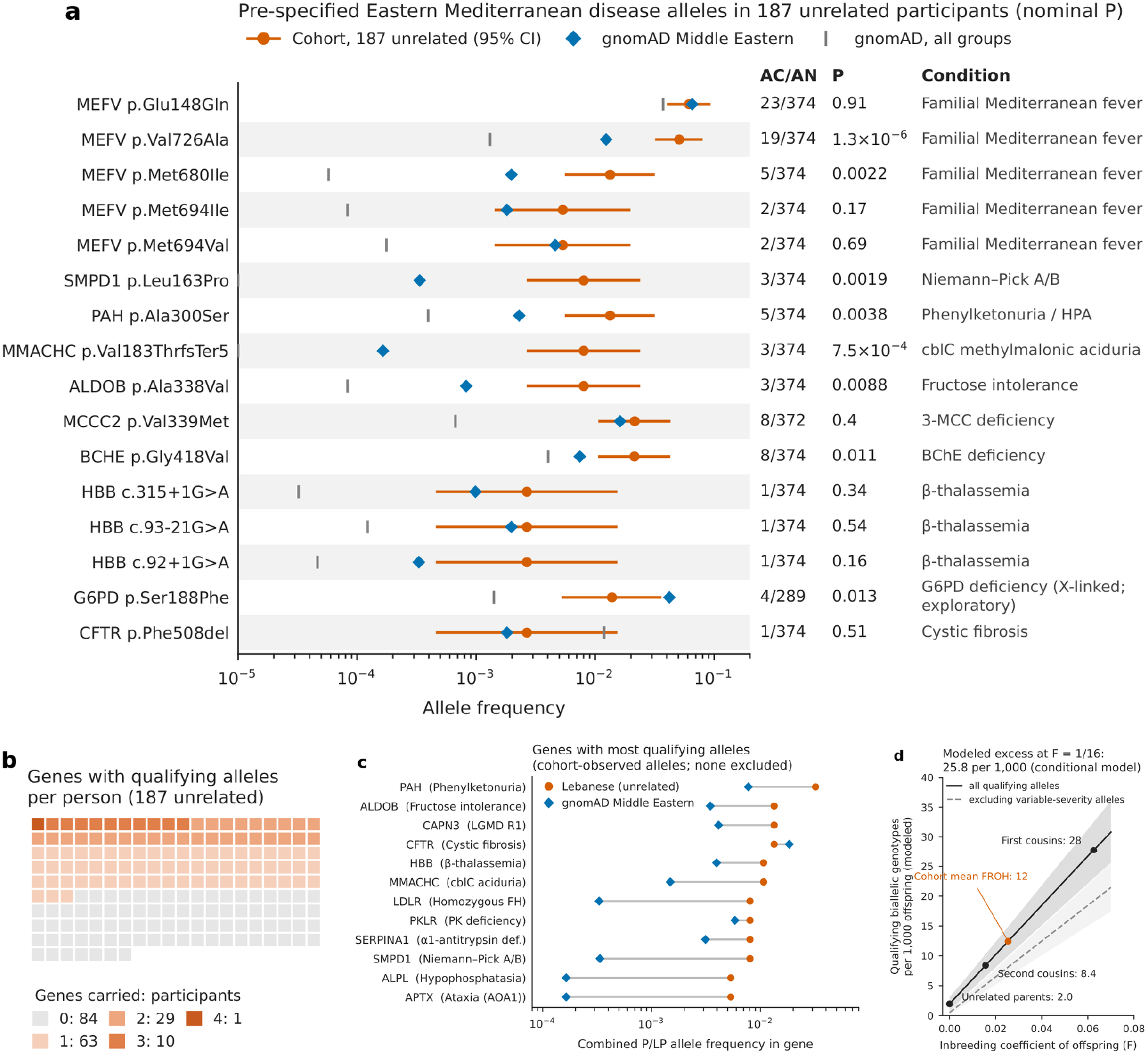
Regional disease alleles and conditional recessive genotype burden. **a**, Pre-specified disease alleles: cohort frequency and Wilson 95% interval (vermilion), MID (blue), and all gnomAD groups (gray). AC/AN denotes cohort allele count/number; P values are nominal Fisher tests. *G6PD* frequencies are per X chromosome. **b**, Qualifying severe-recessive genes per participant; one square per unrelated participant. **c**, Combined frequencies of cohort-observed qualifying alleles in the 12 highest-carrier genes; MID values include the same alleles, not all gene-level carriers. **d**, Modeled qualifying biallelic genotypes per 1,000 offspring versus inbreeding coefficient, before (solid) and after (dashed) severity/inheritance exclusions. Bands are jackknife 95% intervals; the vermilion point marks mean F_ROH_. These are conditional genotypes, not affected births.

### Carrier observations and conditional recessive genotype burden

Using an external list of 1,172 genes for severe, mostly childhood-onset recessive disorders [14–15], 103 of the 187 unrelated participants (55.1%; 95% confidence interval (CI) 47.9–62.0%) carried at least one P/LP allele with at least two ClinVar review stars (Fig. 3b); none was homozygous. Thirty of the 129 qualifying alleles were hypomorphic, of variable severity or disease-causing in heterozygotes, and excluding them left 85 carriers (45.5%; 38.5–52.6%). The genes with most qualifying alleles were *PAH*, *ALDOB*, *CAPN3* and *CFTR* (Fig. 3c). For ten genes, the lower 95% confidence bound of the carrier frequency exceeded 1/200, the American College of Medical Genetics and Genomics (ACMG) carrier-screening threshold [16]; five remained above it after excluding variable-severity alleles (*ALDOB*, *HBB*, *MMACHC*, *PKLR* and *SMPD1*), although these bounds rest on three or four carriers and are fragile (Supplementary Table 12g). The carrier proportion did not increase with autozygosity (58% of participants below F_ROH_ 1/64 versus 52% at or above 1/16).

Under a conditional inbreeding model that pairs distinct observed chromosomes (Materials and Methods), modeled qualifying biallelic genotypes rose from 2.0 per 1,000 offspring of unrelated parents (95% CI 1.1–3.6) to 27.7 per 1,000 offspring of first cousins (23.0–32.5; Fig. 3d), an absolute excess of 25.8 per 1,000 (21.7–29.9), or 18.7 per 1,000 (15.3–22.1) after the exclusions above. Both members of 0.79% of unrelated pairs (0.43–1.44%) carried qualifying alleles in the same gene. These are model outputs conditional on the gene list, the ClinVar release and the alleles seen in 187 people, not predicted affected births.

### Allele-fraction context and candidate inherited-risk findings

Germline allele-fraction filters remove low-fraction somatic calls, so we screened 16 clonal hematopoiesis driver genes in a separate somatic branch and tiered candidates by two criteria: a binomial test that the allele fraction was below 0.5, and an allele fraction at or below the 2.5th percentile of depth- and type-matched singleton heterozygotes from the same callset (223,253 background singletons; Materials and Methods). The screen found 13 driver variants in 12 participants (6.0%; 3.5–10.2%), dominated by *DNMT3A* (five, including p.Arg882His) and truncating *TET2* variants (four), with allele fractions of 0.13 to 0.41 (Fig. 4a).

**Fig 4.**
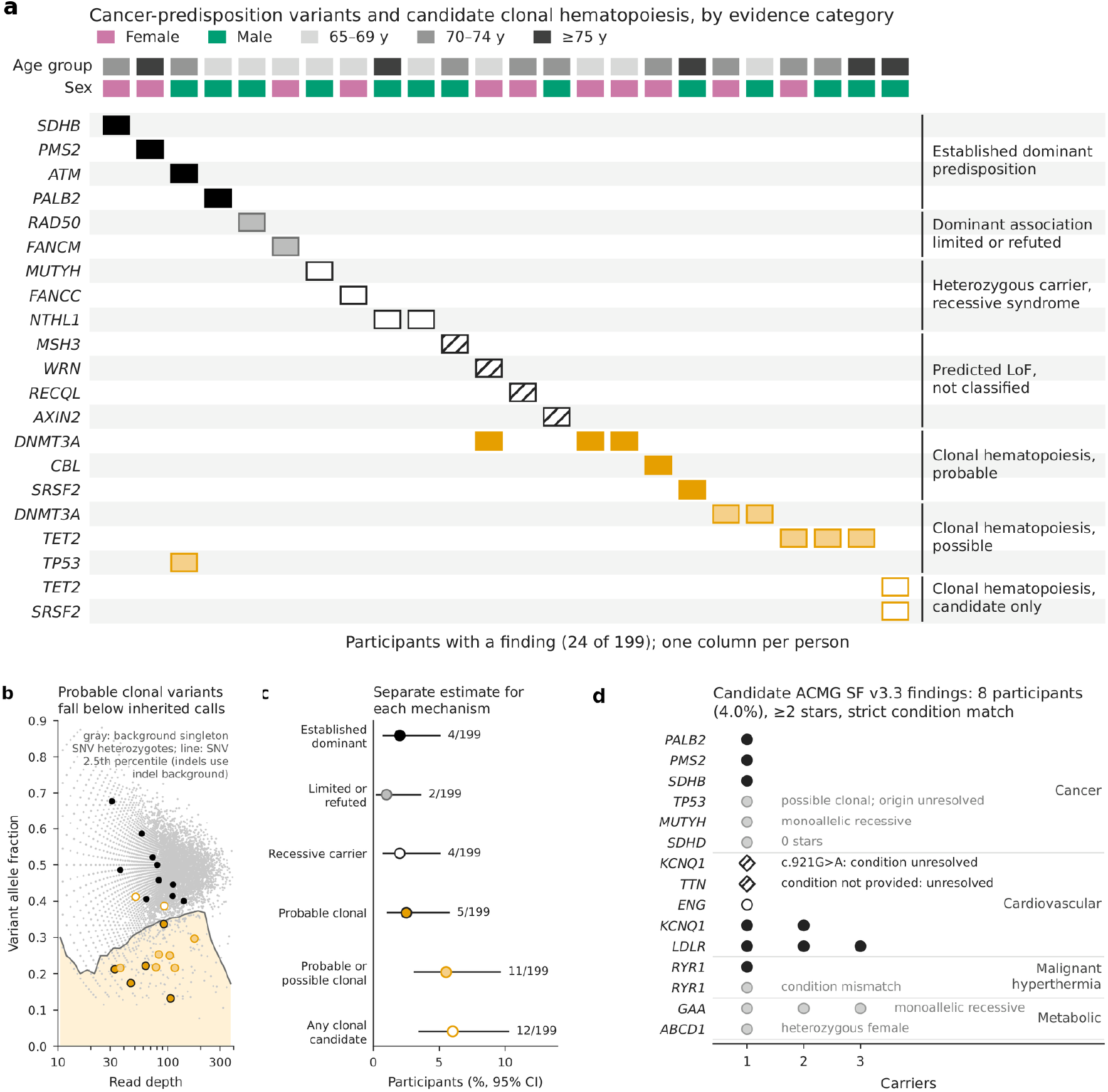
Candidate clonal and inherited-risk findings. **a**, Cancer-panel and clonal-driver variants by participant; rows group inherited-association evidence or clonal tier, with age/sex tracks. **b**, Allele fraction versus depth for putative inherited findings (black) and clonal candidates (probable, dark-edged orange; possible, light orange; candidate only, open). Gray points are presumed-germline singleton SNVs; the curve is their 2.5th percentile. Indels used a separate matched background. **c**, Participant proportions with Wilson 95% intervals. **d**, Secondary-finding candidates by domain: filled circles, ≥2 ClinVar stars and matching condition; open circles, one star; diamonds, unresolved; gray, excluded. No candidate was orthogonally confirmed or clinically adjudicated.

Applied to background singleton heterozygotes presumed to be germline, the binomial criterion flagged 0.8% to 7.8% of SNVs and 1.9% to 17.8% of indels, rising with depth (Supplementary Table 14c). Because these are not confirmed inherited controls, the flagging rates are not validated clinical false-positive rates. Only five participants (2.5%; 1.1–5.7%) met both criteria and were classed as probable clonal hematopoiesis (*DNMT3A* p.Arg882His, p.Gly706AlafsTer72 and p.Lys420Ter, *CBL* p.Cys381Tyr and *SRSF2* p.Pro95His; Fig. 4b). A *TP53* frameshift (allele fraction 0.25; 21 of 83 reads) met the binomial criterion only; because its origin cannot be resolved from blood, it was withheld from germline findings. One man aged 75 or older carried *TET2* p.Arg1134Ter and *SRSF2* p.Pro95Arg at allele fractions of 0.41 and 0.39, compatible with either a large clone or germline variants, and both remain candidates.

After withholding findings of unresolved origin, four participants (2.0%; 0.8–5.1%) carried a P/LP variant in a gene with established dominant cancer predisposition by ClinGen curation (*PALB2*, *PMS2*, *SDHB* and *ATM*; Fig. 4c), two carried truncating variants in genes whose dominant association is limited (*FANCM*) or refuted (*RAD50*), and four were heterozygous for recessive cancer syndromes. Applying ACMG secondary findings (SF) v3.3 reporting rules [17] and requiring the ClinVar condition to match the ACMG condition, eight participants (4.0%; 2.1–7.7%) had candidate secondary findings, in *LDLR* (three), *KCNQ1* (two), and *PALB2*, *PMS2*, *SDHB* and *RYR1* (one each; one woman carried both the *LDLR* and *SDHB* findings; Fig. 4d). Condition matching removed an *RYR1* variant classified for myopathy rather than malignant hyperthermia susceptibility; a *KCNQ1* variant without an explicit long QT syndrome type 1 assertion was classed as unresolved (nine participants, 4.5%, if included).

None of the cancer-predisposition or secondary-finding candidates was orthogonally confirmed or clinically adjudicated.

## Discussion

The genetic interpretation of these exomes depended on distinguishing quantities that can otherwise be conflated. Overall absence from a reference did not behave like the frequency of particular regional alleles; the proportion carrying a qualifying recessive allele differed from the expected number of biallelic genotypes; and a blood-detected driver variant did not necessarily identify either a constitutional allele or a confirmed clone. Together, these observations connect population sampling, shared ancestry and tissue-specific variation without reducing them to a single measure of underrepresentation.

The reference comparison provides the clearest example of how the statistical benchmark changed the interpretation. Unmatched absence was much higher than a beta-binomial expectation, but that expectation also underestimated absence between cohort subsets. At matched allele number, the reference-to-cohort absence ratio for single-nucleotide variants was close to one. Thus, a large raw absence fraction did not establish a corresponding excess of cohort-specific exome variants. The relevant observation was conditional: for alleles detected by this exome workflow, a size-matched Middle Eastern reference sample missed them at approximately the rate of a similarly sized cohort comparison. The overlapping splits describe robustness to partitioning within this dataset, not independent replication across populations.

This result does not argue against expanding regional genomic resources. It leaves structural variants, difficult repetitive regions and most noncoding sequence untested. Long-read assembly studies identifying additional Middle Eastern sequence address complementary gaps in representation and detection [18]. Even among observed exome alleles, pooled reference groups can obscure local frequencies. Reference-size matching and geographically informative sampling therefore address different problems: one checks an absence benchmark, while the other supplies allele observations and population structure that a pooled resource may not capture.

The recurrent reference-absent alleles must also be distinguished from the disease-allele panel. None of the 360 recurrent alleles absent from full MID was ClinVar pathogenic or likely pathogenic. The nominally enriched *MEFV*, *SMPD1* and other regional disease alleles arose from a separate pre-specified comparison; they were not shown to be members of that absent set. *MEFV* founder haplotypes have been reported in Lebanon, and the regional familial Mediterranean fever spectrum is established [7,19]. Our findings document frequency differences in these selected older participants, rather than discover a new disease mechanism or establish pathogenicity through enrichment. The candidate-panel comparisons are nominal, and *MEFV* p.Val726Ala did not cross the genome-wide threshold.

Autozygosity provides a second connection between demographic context and genotype interpretation. The carrier proportion did not rise with autozygosity, yet the conditional model predicted a substantial increase in qualifying biallelic genotypes at first-cousin inbreeding coefficients. This is not contradictory: carrying one qualifying copy and pairing two related copies are different events. The estimate is sensitive to the allele definition, as shown by the reduction after severity-related exclusions. The model is conditional on the observed alleles, gene list and ClinVar release, and does not incorporate complete penetrance or genotype-specific severity. It must not be read as a prediction of affected births.

The regional carrier observations are consequently more informative for allele-level context than for a new screening policy. Genes such as *SMPD1* and *MMACHC* already appear in carrier-screening guidance, and broader population analyses include alleles not seen here [16,20]. Studies of recessive pathogenic variation differ in population, ascertainment and qualifying criteria [21–22], making unadjusted comparisons of carrier percentages difficult. Lower confidence bounds above a carrier-frequency threshold are also fragile when supported by only three or four individuals. A geographically broader and younger sample would be needed to estimate reproductive-population frequencies more precisely.

The blood-derived candidates illustrate a third distinction: a plausible biological annotation is not equivalent to evidence of origin. A binomial imbalance test flagged a depth-dependent fraction of background singletons, indicating that the theoretical heterozygous expectation did not capture all dispersion in this callset. Those singletons are presumed germline, not confirmed inherited controls, so their flagging rates are not validated clinical false-positive rates. Adding an empirical tail criterion narrowed the probable tier but did not prove clonal origin. Conversely, a candidate that failed that criterion could still be acquired. Keeping the *TP53* frameshift unresolved was therefore more defensible than assigning it to an inherited-risk category solely from blood, consistent with the wider ambiguity of blood-detected *TP53* findings [10].

Condition matching further separated annotation from interpretation. A myopathy-associated *RYR1* classification did not establish the secondary-finding condition of malignant hyperthermia susceptibility. Similarly, the 4.0% secondary-finding proportion describes unconfirmed research candidates, not a demonstrated actionable yield. Lebanese diagnostic cohorts and validated return-of-results programs elsewhere provide relevant but non-equivalent comparisons [23–25]. Technical confirmation, variant-origin assessment when indicated and clinical adjudication would be necessary before individual return-of-results decisions.

The cohort and assay constrain all of these conclusions. Participants were selected for an older ambulatory vitamin D trial with important disease exclusions, and DNA availability and budget determined exome inclusion. The data do not represent all Lebanese communities or support penetrance estimation. Frequency estimates rely on only 187 unrelated participants; missing genotypes counted as non-carriers make proportions detection proportions. Exome-marker spacing affects homozygosity estimates, while moderate-depth exomes and a germline caller limit sensitivity for small clones. There was no independent cohort validation, orthogonal confirmation or non-blood testing, and severity flags require continued curation.

In summary, these population exomes were most informative when reference sampling, local frequencies, autozygosity and possible acquired variation were evaluated separately. Size matching altered the interpretation of bulk reference absence without erasing regional allele differences. Shared ancestry changed conditional genotype expectations without establishing disease incidence, and allele-fraction context supported cautious candidate tiering without resolving tissue of origin. Independent datasets and appropriate validation are needed to determine how far these observations extend beyond this cohort.

## Materials and Methods

### Ethics statement

The parent trial and prior exome report document written informed consent and approval by the American University of Beirut Institutional Review Board (IM-GEHF-20, 24 January 2011). The parent trial is registered as NCT01315366.

### Participants and study design

This is a secondary observational analysis of previously generated trial exomes. Participants were enrolled between January 2011 and July 2013 in a double-blind trial of calcium with two doses of vitamin D_3_ in greater Beirut (NCT01315366) [12]. Inclusion required an age of at least 65 years, a body mass index above 25 kg m^−2^, ambulatory status and a serum 25-hydroxyvitamin D concentration of 10–30 ng ml^−1^ at screening. People with diabetes, cancer, autoimmune disease, severe cardiac, hepatic or renal disease, or conditions affecting bone metabolism were excluded [12]. Exome data were available for 199 of the 257 randomized participants [11]; selection was limited by the availability and quality of DNA and by budget.

### Participants were recruited at three Beirut hospitals (145 at the American University of Beirut

Medical Center, 51 at Hôtel-Dieu de France and 3 at Rafik Hariri University Hospital). They constitute a selected group of ambulatory trial participants rather than a population-representative sample, and genotype frequencies were not used to infer penetrance. The trial report [12] and the exome study [11] report written informed consent and approval by the American University of Beirut Institutional Review Board (IM-GEHF-20, 24 January 2011); the trial is registered as NCT01315366.

### Sequencing and variant calling

Peripheral blood DNA was extracted with the QIAamp DNA Blood Midi Kit (QIAGEN). Exome libraries were prepared with the Agilent SureSelectXT Human All Exon V5 design (S04380110) and sequenced by Macrogen on an Illumina HiSeq 4000 (101-bp paired-end reads; 68.8 to 91.8 million reads and 6.9 to 9.2 Gb per sample; 95.9% to 96.7% of bases with Phred quality ≥30) [11]. For the present analysis, reads were mapped to the GRCh38 primary assembly with CLC Genomics Workbench 26.0 (QIAGEN). Duplicates were marked, and base-quality scores were recalibrated with GATK 4.6.2.0 using dbSNP 157, known indels, and the Mills and 1000 Genomes gold-standard indels. GATK HaplotypeCaller was run per sample in GVCF mode, with chromosomes X and Y processed separately, and GVCFs were joint-genotyped with GenomicsDBImport and GenotypeGVCFs. Multiallelic sites were decomposed and normalized with bcftools 1.23.1 ( [26]) with reference-allele checking, calls were restricted to the capture targets with 100-bp padding, and separate GATK hard filters were applied to single-nucleotide variants (SNVs) (QD < 2, QUAL < 30, FS > 60, SOR > 3, MQ < 40, MQRankSum < −12.5, ReadPosRankSum < −8) and indels (QD < 2, QUAL < 30, FS > 200, ReadPosRankSum < −20), retaining PASS variants. All genotypes in the joint-called VCF are encoded as diploid; X-chromosome ploidy was assigned in the analysis from genetic sex.

### Sample quality control and genotype filtering

Call rate, mean depth, transition/transversion ratio, heterozygous/homozygous ratio, X-chromosome heterozygosity and allele-balance dispersion were computed from the joint-called VCF (Supplementary Table 17 and Supplementary Fig. 13). Transition/transversion ratios ranged from 2.33 to 2.41, and allele-balance dispersion identified no contamination outliers. The source study excluded five samples with call rates below 95% because of off-target flanking sequence [11]; because the present callset is restricted to the capture targets, all 199 samples were retained, and neither of the two samples below 95% here is among those five. Recorded and genetic sex disagreed for three participants; genetic sex was used for them, and none carried a cancer-predisposition, clonal or secondary finding. Headline proportions changed by at most one percentage point after excluding the low-call-rate and sex-discordant samples, singly or together (Supplementary Table 18).

Every non-reference genotype was required to have depth ≥10, at least three alternate reads, and an alternate-read fraction ≥0.20 for heterozygotes or ≥0.80 for homozygous-alternate calls; failing genotypes were masked. Sites with GATK ExcessHet above 54.69, the threshold of the GATK joint-genotyping workflow [27–28], were excluded. Allele numbers were counted from callable genotypes (depth ≥10). For carrier proportions, masked or missing genotypes were counted as non-carriers. Callable allele numbers for headline alleles are given in Supplementary Table 21b, and the unfiltered baseline in Supplementary Figs. 3–5 and 11.

### Ancestry, relatedness and autozygosity

Principal components were computed in PLINK v2.0.0-a.6.9LM [29] jointly with 3,280 HGDP and 1000 Genomes samples [30] on 3,176 LD-pruned markers; the first three components explained 35.7%, 31.8% and 10.6% of the variance captured by the first ten. Relatedness was estimated as KING-robust kinship [31] on 32,622 LD-pruned markers (*r*^2^ < 0.2, 500-kb windows). Fifteen pairs had kinship above 0.0442 (8 first-degree, 1 second-degree and 6 third-degree); removing one member of each left 187 unrelated participants, the denominator for frequency contrasts, absence calibration and recessive carriers. Cancer, clonal hematopoiesis, secondary-finding and *MEFV* carrier proportions use all 199. Runs of homozygosity were called with bcftools roh [26,32] on 96,978 high-quality common SNVs; runs of at least 1.5 Mb, 25 markers and quality 30 were summed and divided by 2,875 Mb of autosomal sequence to give F_ROH_ ( [33]). A SNP-based inbreeding coefficient was computed from LD-pruned markers, and estimates were repeated with gap splitting, alternative marker sets, long runs only and MID-based allele frequencies (Supplementary Table 13). Participants were also classified by total-ROH thresholds derived from pedigree-confirmed Lebanese exomes [34], giving 44 (22.1%) consanguineous and 18 (9.0%) probably consanguineous participants (Supplementary Table 13b).

### Comparison with the Middle Eastern reference

Alleles were matched to gnomAD v4.1 joint exome and genome data [2,35] and classified as observed in MID, not observed in MID with a positive MID allele number (callable absence), recorded with a zero MID allele number, or lacking a MID record (Supplementary Table 20); only callable absence was interpreted. Allelic Fisher exact tests compared allele counts in the 187 unrelated participants with MID for 93,724 coding or splice variants at a Bonferroni threshold of 0.05/93,724 (5.3 × 10^−7^); results for all 199 participants are in Supplementary Tables 2 and 2b. For a pre-specified set of Eastern Mediterranean disease alleles (Supplementary Table 15), nominal Fisher *P* values are reported without multiplicity correction. For X-linked alleles, male genotypes contributed one allele, and heterozygous calls in men at non-pseudoautosomal sites were treated as missing.

### Size-matched absence calibration

Calibration used autosomal sites only, where each person contributes two allele copies. The 187 unrelated participants were split at random into halves of 94 and 93 people ten times (seed 11). For each allele seen in the first (discovery) half, we recorded whether it was absent from the 186 allele copies of the 93 people in the second half, and computed the probability that it would be absent from 186 allele copies drawn at random from MID (hypergeometric, from the MID allele count and allele number). The two absence rates were compared by the number of copies in the discovery half, by variant type, and among sites callable (depth ≥10) in at least 189 of the 199 participants or in every member of the comparison half (Supplementary Tables 21 and 21c). The overlapping splits are not independent replicates; ranges across splits are descriptive, not confidence intervals from independent cohorts. For comparison, we computed beta-binomial expectations with a Jeffreys prior.

### Clinical annotation and cancer-predisposition genes

Variants were annotated with OpenCRAVAT 3.1.1 ( [36]) and the July 2026 ClinVar release [37]; ClinVar records for secondary-finding candidates were re-queried on 28 September 2026.

ClinVar pathogenic or likely pathogenic (P/LP) variants with at least one review star were evaluated. Variants in 75 cancer-predisposition genes were assigned, using ClinGen gene–disease validity curations (download of 28 September 2026) [38–39], to established dominant predisposition (a Definitive or Strong autosomal-dominant cancer curation), dominant association limited or refuted, heterozygous carrier of a recessive cancer syndrome, or predicted loss of function without a ClinVar P/LP classification (Supplementary Table 19). Proportions are reported with Wilson 95% intervals [40].

### Clonal hematopoiesis

We screened *DNMT3A*, *TET2*, *ASXL1*, *PPM1D*, *TP53* and *BRCC3* for truncating variants, and *DNMT3A*, *JAK2*, *SF3B1*, *SRSF2*, *U2AF1*, *IDH1*, *IDH2*, *GNB1*, *KRAS*, *NRAS* and *CBL* for established hotspots [8–9]. This screen used a separate somatic branch (depth ≥10, at least three alternate reads, no allele-fraction floor). Candidates meeting both a one-sided binomial test that the allele fraction was below 0.5 (*P* < 0.01) and an allele fraction at or below the 2.5th percentile of depth- and type-matched singleton heterozygotes from the same callset (223,253 background singletons, presumed germline rather than confirmed inherited controls) were classed as probable, those meeting only the binomial criterion as possible, and the remainder as candidates only. The binomial criterion was assessed by applying it to the background singletons (Supplementary Table 14c). Artifacts were assessed from alternate reads at the same position in the other 198 participants and from strand-bias and mapping-quality metrics; none of the 13 positions carried a single alternate read in any other participant (Supplementary Tables 14 and 14b). Candidates by age group are shown in Supplementary Fig. 15.

### Recessive genotypes and inbreeding model

Genes for severe autosomal-recessive disorders were taken from the Mackenzie’s Mission reproductive carrier-screening list [14] (PanelApp Australia panel 3139 v1.0; 1,172 green biallelic autosomal genes) [15]. For each unrelated participant we counted genes carrying a filtered P/LP allele with at least two ClinVar review stars, excluding clonal hematopoiesis genes. Each qualifying allele was mapped to its condition, and alleles that are hypomorphic, of variable severity or disease-causing in heterozygotes were flagged and excluded in a sensitivity analysis (Supplementary Table 12e); this flagging is provisional. Gene-level carrier frequencies, with Wilson intervals, were compared with the ACMG 1/200 threshold [16] (Supplementary Table 12g). For gene *g* with *K∼g∼* qualifying alleles among *M* = 374 chromosomes, the probability that two distinct chromosomes both carry a qualifying allele was estimated as *q∼g∼* = *K∼g∼*(*K∼g∼* − 1)/[*M*(*M* − 1)], avoiding the self-pairing bias of the plug-in estimator. The expected number of qualifying biallelic genotypes per offspring at inbreeding coefficient *F* was *E*(*F*) = Σ_g_ [(1 − *F*)*q∼g∼* + *F K∼g∼*/*M*]. This sum approximates the probability of at least one qualifying genotype only when multiple qualifying genotypes in one offspring are uncommon. The carrier-pair frequency was the proportion of pairs of distinct participants carrying qualifying alleles in the same gene. Intervals are delete-one jackknife 95% intervals (Supplementary Table 12f). Assumptions, including phase and the pairing of people rather than chromosomes, are given in the Supplementary Methods.

### Secondary findings

Secondary-finding review evaluated P/LP variants in the 84 genes of ACMG SF v3.3 ( [17]). The primary estimate required at least two ClinVar review stars; one-star and unresolved matches were displayed separately. Single heterozygous variants in *GAA* and *MUTYH*, and heterozygous *ABCD1* variants in women, were not counted. Variants with zero review stars or classed as probable clonal hematopoiesis were excluded, and findings of unresolved somatic or constitutional origin were withheld. The ClinVar condition was required to match the ACMG SF condition for the gene, and *TTN* was restricted to truncating variants; matches needing curation were reported as unresolved. None has been confirmed orthogonally or adjudicated clinically (Supplementary Tables 5 and 16).

### Statistics

Allelic comparisons used two-sided Fisher exact tests; proportions are reported with Wilson 95% intervals and modeled genotype burdens with jackknife intervals. Correlations used Pearson’s *r* and Spearman’s *ρ*. Analyses used cyvcf2 0.34.0 and Python 3.12 (NumPy 2.5.3, pandas 3.0.6, SciPy 1.18.1, statsmodels 0.15.0 and matplotlib).

## Data Availability

Aggregate allele-frequency tables, numerical figure source data, analysis metadata and code are intended to be shared with the article or through versioned repositories. Individual-level blood-derived BAM, GVCF and joint-called VCF data are not publicly released because of participant privacy. Analysis code covers quality control, genotype filtering, autozygosity, reference calibration, clonal-candidate tiering, recessive modeling, secondary-finding review and figures. No new repository deposition, accession or approval is asserted in this draft.

## Acknowledgments

This work was partly performed using the American University of Beirut High-Performance Computing cluster Octopus, made available through the Research Computing Center under the School of Computing and Data Science.

## AI-assisted work

As reported in the source manuscript, Claude (Anthropic) was used under author direction for analysis code, data figures, literature assistance, reference checking and text revision. Biomni (Phylo) assisted with the present journal-specific restructuring, language editing and document preparation; no new genomic analyses were performed during this revision.

## Author contributions

N.E.-H. conceived the study, designed and performed the computational genomic analyses and wrote the manuscript. G.N. provided the raw sequencing data and BAM files, oversaw sequencing, contributed to sample collection and reviewed the manuscript. N.N.H. and G.E.-H.F. contributed to sample collection and previous analyses of the cohort and reviewed the manuscript. M.E.-H. contributed to data analysis and writing and reviewing the manuscript.

## References

1. Bhattacharya R, Chen N, Shim I, Kuwahara H, Gao X, Alkuraya FS, et al. Massive underrepresentation of Arabs in genomic studies of common disease. Genome Med. 2023;15:99. 10.1186/s13073-023-01254-8

2. gnomAD Production Team. gnomAD v4.1 release announcement. https://gnomad.broadinstitute.org/news/2024-04-gnomad-v4-1. 2024. Accessed 28 Oct 2026.

3. Richards S, Aziz N, Bale S, Bick D, Das S, Gastier-Foster J, et al. Standards and guidelines for the interpretation of sequence variants: a joint consensus recommendation of the American College of Medical Genetics and Genomics and the Association for Molecular Pathology. Genet Med. 2015;17:405–424. 10.1038/gim.2015.30

4. Whiffin N, Minikel E, Walsh R, O’Donnell-Luria AH, Karczewski K, Ing AY, et al. Using high-resolution variant frequencies to empower clinical genome interpretation. Genet Med. 2017;19:1151–1158. 10.1038/gim.2017.26

5. Barbour B, Salameh P. Consanguinity in Lebanon: prevalence, distribution and determinants. J Biosoc Sci. 2009;41:505–517. 10.1017/s0021932009003290

6. Jalkh N, Sahbatou M, Chouery E, Megarbane A, Leutenegger AL, Serre JL. Genome-wide inbreeding estimation within Lebanese communities using SNP arrays. Eur J Hum Genet. 2015;23:1364–1369. 10.1038/ejhg.2014.246

7. Jalkh N, Génin E, Chouery E, Delague V, Medlej-Hashim M, Idrac CA, et al. Familial Mediterranean Fever in Lebanon: founder effects for different MEFV mutations. Ann Hum Genet. 2008;72:41–47. 10.1111/j.1469-1809.2007.00386.x

8. Jaiswal S, Fontanillas P, Flannick J, Manning A, Grauman PV, Mar BG, et al. Age-related clonal hematopoiesis associated with adverse outcomes. N Engl J Med. 2014;371:2488–2498. 10.1056/nejmoa1408617

9. Genovese G, Kähler AK, Handsaker RE, Lindberg J, Rose SA, Bakhoum SF, et al. Clonal hematopoiesis and blood-cancer risk inferred from blood DNA sequence. N Engl J Med. 2014;371:2477–2487. 10.1056/nejmoa1409405

10. MacGregor HAJ, Blundell JR, Easton DF. Shared inheritance reveals landscape of somatic and germline cancer risk in TP53. Am J Hum Genet. 2026;113:1916–1928. 10.1016/j.ajhg.2026.07.011

11. Hendi NN, Chakhtoura M, Al-Sarraj Y, Basha DS, Albagha O, Fuleihan GE, et al. The Genetic Architecture of Vitamin D Deficiency among an Elderly Lebanese Middle Eastern Population: An Exome-Wide Association Study. Nutrients. 2023;15:3216. 10.3390/nu15143216

12. Rahme M, Sharara SL, Baddoura R, Habib RH, Halaby G, Arabi A, et al. Impact of Calcium and Two Doses of Vitamin D on Bone Metabolism in the Elderly: A Randomized Controlled Trial. J Bone Miner Res. 2017;32:1486–1495. 10.1002/jbmr.3122

13. Mancuso DJ, Tuley EA, Westfield LA, Lester-Mancuso TL, Le Beau MM, Sorace JM, et al. Human von Willebrand factor gene and pseudogene: structural analysis and differentiation by polymerase chain reaction. Biochemistry. 1991;30:253–269. 10.1021/bi00215a036

14. Kirk EP, Ong R, Boggs K, Hardy T, Righetti S, Kamien B, et al. Gene selection for the Australian Reproductive Genetic Carrier Screening Project (“Mackenzie’s Mission”). Eur J Hum Genet. 2021;29:79–87. 10.1038/s41431-020-0685-x

15. PanelApp Australia. Mackenzie’s Mission gene list, panel 3139, version 1.0. https://panelapp-aus.org/panels/3139/. 2026. Accessed 28 Oct 2026.

16. Gregg AR, Aarabi M, Klugman S, Leach NT, Bashford MT, Goldwaser T, et al. Screening for autosomal recessive and X-linked conditions during pregnancy and preconception: a practice resource of the American College of Medical Genetics and Genomics (ACMG). Genet Med. 2021;23:1793–1806. 10.1038/s41436-021-01203-z

17. Lee K, Abul-Husn NS, Amendola LM, Brothers KB, Chung WK, Gollob MH, et al. ACMG SF v3.3 list for reporting of secondary findings in clinical exome and genome sequencing: A policy statement of the American College of Medical Genetics and Genomics (ACMG). Genet Med. 2025;27:101454. 10.1016/j.gim.2025.101454

18. Ghorbani M, Moosa S, Siddig Z, Farhad R, Naeem H, Harvey WT, et al. Near-complete Middle Eastern genomes refine autozygosity and enhance disease-causing and population-specific variant discovery. Nat Genet. 2025;57:1119–1131. 10.1038/s41588-025-02173-7

19. Touitou I. The spectrum of Familial Mediterranean Fever (FMF) mutations. Eur J Hum Genet. 2001;9:473–483. 10.1038/sj.ejhg.5200658

20. Schmitz MJ, Bashar A, Soman V, Nkrumah EAF, Al Mulla H, Darabi H, et al. Leveraging diverse genomic data to guide equitable carrier screening: Insights from gnomAD v.4.1.0. Am J Hum Genet. 2025;112:181-195. 10.1016/j.ajhg.2024.11.004

21. Fridman H, Yntema HG, Mägi R, Andreson R, Metspalu A, Mezzavila M, et al. The landscape of autosomal-recessive pathogenic variants in European populations reveals phenotype-specific effects. Am J Hum Genet. 2021;108:608–619. 10.1016/j.ajhg.2021.03.004

22. Aamer W, Al-Maraghi A, Syed N, Gandhi GD, Aliyev E, Al-Kurbi AA, et al. Burden of Mendelian disorders in a large Middle Eastern biobank. Genome Med. 2024;16:46. 10.1186/s13073-024-01307-6

23. Jalkh N, Mehawej C, Chouery E. Actionable Exomic Secondary Findings in 280 Lebanese Participants. Front Genet. 2020;11:208. 10.3389/fgene.2020.00208

24. Hanna EM, Mehawej C, Hoblos Y, Rahy K, Megarbane A, Chouery E. Certain vs. uncertain actionable secondary findings in a cohort of 500 Lebanese participants: What to report to the patient?. PLoS One. 2025;20:e0327471. 10.1371/journal.pone.0327471

25. Kerr SM, Klaric L, Muckian MD, Johnston K, Drake C, Halachev M, et al. Actionable genetic variants in 4,198 Scottish participants from the Orkney and Shetland founder populations and implementation of return of results. Am J Hum Genet. 2025;112:793–807. 10.1016/j.ajhg.2025.02.018

26. Danecek P, Bonfield JK, Liddle J, Marshall J, Ohan V, Pollard MO, et al. Twelve years of SAMtools and BCFtools. Gigascience. 2021;10:giab008. 10.1093/gigascience/giab008

27. Broad Institute. GATK tool documentation: ExcessHet. https://gatk.broadinstitute.org/hc/en-us/articles/360037261311-ExcessHet. 2026. Accessed 28 Oct 2026.

28. Broad Institute. WARP JointGenotyping pipeline overview. https://broadinstitute.github.io/warp/docs/Pipelines/JointGenotyping_Pipeline/README. 2026. Accessed 28 Oct 2026.

29. Chang CC, Chow CC, Tellier LC, Vattikuti S, Purcell SM, Lee JJ. Second-generation PLINK: rising to the challenge of larger and richer datasets. Gigascience. 2015;4:7. 10.1186/s13742-015-0047-8

30. Koenig Z, Yohannes MT, Nkambule LL, Zhao X, Goodrich JK, Kim HA, et al. A harmonized public resource of deeply sequenced diverse human genomes. Genome Res. 2024;34:796–809. 10.1101/gr.278378.123

31. Manichaikul A, Mychaleckyj JC, Rich SS, Daly K, Sale M, Chen WM. Robust relationship inference in genome-wide association studies. Bioinformatics. 2010;26:2867–2873. 10.1093/bioinformatics/btq559

32. Narasimhan V, Danecek P, Scally A, Xue Y, Tyler-Smith C, Durbin R. BCFtools/RoH: a hidden Markov model approach for detecting autozygosity from next-generation sequencing data. Bioinformatics. 2016;32:1749–1751. 10.1093/bioinformatics/btw044

33. McQuillan R, Leutenegger AL, Abdel-Rahman R, Franklin CS, Pericic M, Barac-Lauc L, et al. Runs of homozygosity in European populations. Am J Hum Genet. 2008;83:359–372. 10.1016/j.ajhg.2008.08.007

34. Hanna EM, Mehawej C, Assy J, Corbani S, Korban R, Megarbane A, et al. Predicting Consanguinity Rates from Exome Sequencing Data in the Lebanese Population. J Mol Diagn. 2025;27:177–183. 10.1016/j.jmoldx.2024.11.008

35. Karczewski KJ, Francioli LC, Tiao G, Cummings BB, Alföldi J, Wang Q, et al. The mutational constraint spectrum quantified from variation in 141,456 humans. Nature. 2020;581:434–443. 10.1038/s41586-020-2308-7

36. Pagel KA, Kim R, Moad K, Busby B, Zheng L, Tokheim C, et al. Integrated Informatics Analysis of Cancer-Related Variants. JCO Clin Cancer Inform. 2020;4:310–317. 10.1200/cci.19.00132

37. Landrum MJ, Lee JM, Benson M, Brown GR, Chao C, Chitipiralla S, et al. ClinVar: improving access to variant interpretations and supporting evidence. Nucleic Acids Res. 2018;46:D1062–D1067. 10.1093/nar/gkx1153

38. Strande NT, Riggs ER, Buchanan AH, Ceyhan-Birsoy O, DiStefano M, Dwight SS, et al. Evaluating the Clinical Validity of Gene-Disease Associations: An Evidence-Based Framework Developed by the Clinical Genome Resource. Am J Hum Genet. 2017;100:895–906. 10.1016/j.ajhg.2017.04.015

39. ClinGen. Gene–disease validity curations. https://search.clinicalgenome.org/kb/gene-validity. 2026. Accessed 28 Oct 2026.

40. Wilson EB. Probable inference, the law of succession, and statistical inference. J. Am. Stat. Assoc. 1927;22:209–212.

