## Supplemental methods and figures for "Reference calibration, autozygosity and blood-derived variant interpretation in older adults"

### Supplementary Information

#### Founder alleles and clonal hematopoiesis, not missing diversity, confound variant interpretation in an underrepresented population

Contents: Supplementary Methods; Figures S1 to S14; list of Supplementary Tables S1 to S21, which are supplied as one Excel workbook, with large tables also as compressed CSV; Supplementary references.

### Supplementary Methods

#### Source cohort and callset

The participants were enrolled in a randomized trial of calcium with two doses of vitamin D3 in greater Beirut (NCT01315366; January 2011 to July 2013) [1, 2]. The source exome study sequenced 199 participants and analyzed 194 after excluding five with call rates below 95%; its published workflow aligned reads with BWA-MEM to GRCh37 [1]; the present analysis remapped the reads to GRCh38. Processing for the present analysis (as documented by the authors): FASTQ files were mapped to the GRCh38 primary assembly with CLC Genomics Workbench 26.0 (BAM header @PG PN:clcgenomicswb VN:26.0); BAM processing, per-sample calling and joint genotyping, which produced the GVCFs and the VCF used for all results, were done with GATK 4.6.2.0 on the AUB Octopus cluster (duplicate marking; base-quality recalibration with dbSNP 157, known indels and the Mills and 1000 Genomes gold-standard indels); GATK HaplotypeCaller was run per sample in GVCF mode with chromosomes X and Y processed separately; and the GVCFs were joint-genotyped with GenomicsDBImport and GenotypeGVCFs. Verified against the header of the supplied VCF (cohort.v5_pad100.hard.PASS.vcf.gz; SHA-256 f38d6462ec7f0a96a0b3ff71623235a1d4dcaf4421524dc8ae1c22705df6d8cb): GATK 4.6.2.0 GenomicsDBImport (30 July 2026; scattered intervals, batch size 50); FILTER definitions for the GATK hard filters, applied separately to SNVs (QD2, QUAL30, FS60, SOR3, MQ40, MQRankSum-12.5 and ReadPosRankSum-8; that is, QD <2, QUAL <30, FS >60, SOR >3, MQ <40, MQRankSum <−12.5, ReadPosRankSum <−8) and to indels (QD2, QUAL30, FS200 and ReadPosRankSum-20; QD <2, QUAL <30, FS >200, ReadPosRankSum <−20). The assignment was confirmed on the PASS records: no SNV exceeds any SNV threshold, whereas 395 PASS indels have FS >60 and 4,295 have SOR >3, and no indel exceeds FS 200 or falls below ReadPosRankSum −20; per-chromosome concatenation and left-normalization with splitting of multiallelic records by bcftools 1.23.1 against Homo_sapiens.GRCh38.dna.primary_assembly.fa with –check-ref e, which stops on any reference-allele mismatch; and restriction to the capture-target file S04380110_Covered (SureSelect Human All Exon V5) padded by 100 bp. The file has 25 contigs without the chr prefix and no ##reference line. Checked on the current file: the record, genotype and allele-count recount (below); exact matching of 96.5% of filtered alleles to a gnomAD v4.1 GRCh38 record by position, reference and alternate allele; per-sample transition/transversion ratios of 2.33 to 2.41; and genotype ploidy. All genotypes, including those on non-pseudoautosomal X and on Y, are encoded as diploid, so X ploidy was assigned in the analysis from genetic sex, and heterozygous calls at non-pseudoautosomal X sites in males are reported (Figure S8). The five samples excluded by the source study had low call rates there because off-target flanking sequence was retained; restricting calls to the targeted regions removes it, and neither of the two samples below 95% in the present callset is among those five. It contains 199 sample columns and 656,024 records (570,139 SNVs and 85,885 indels or complex records). One record (1:207063889:G:T in PFKFB2) has AC 0 and AN 396; it is counted as a record but not as an observed alternate allele, so observed-allele denominators are 656,023. No multiallelic records, symbolic alleles or partial genotypes were present. No further realignment or recalling was done in the downstream analysis.

#### Sample quality control

Per-sample metrics were computed from the supplied callset (Table S17, Figure S13): - autosomal SNV call rate; - mean genotype depth; - fraction of sites with depth ≥10; - transition/transversion ratio; - heterozygous to homozygous-alternate ratio; - non-pseudoautosomal X heterozygosity; - fraction of heterozygous genotypes with allele balance outside 0.3–0.7, as a contamination proxy.

Samples are listed under anonymous codes ordered by call rate. Headline analyses were repeated after excluding (i) the two samples with call rates below 95% (Q001, Q002), (ii) the five lowest-call-rate samples (Q001–Q005; call rates 92.9–95.3%), (iii) the three sex-discordant samples (Q117, Q141, Q155), and (iv) sets (ii) and (iii) together (eight samples; no overlap) (Table S18). These sets are defined from the present callset; they differ from the five samples excluded by the source study (see above). Table S18 uses the current endpoint definitions (probable clonal hematopoiesis requires both allele-fraction criteria; secondary findings use the strict primary rule). None of the three sex-discordant participants carried a cancer-predisposition, clonal hematopoiesis, secondary-finding or G6PD allele; one carried a classic MEFV allele.

#### Genotype recount and read-support rules

Records were decoded genotype by genotype. AC was the number of alternate alleles and AN the number of called alleles in complete genotypes. The recount reproduced AC, AN, called, missing, heterozygous and homozygous-alternate counts for all 656,024 records of the supplied summary table.

The read-support rules were those stated for the callset: - site PASS and QUAL ≥20; - genotype depth ≥10; - for non-reference genotypes, at least three alternate reads and an alternate-read fraction of at least 0.20 for heterozygous or 0.80 for homozygous-alternate calls.

Of the 18,245,299 non-reference genotypes, 2,813,639 (15.4%) failed at least one rule; 81% of failures occurred at sites with mean depth below 15 (Figure S3). Failing genotypes were masked. Allele numbers were taken from callable genotypes: depth

≥10, or a non-reference call that met the rules. For carrier proportions, masked or missing genotypes were counted as non-carriers.

#### Excess heterozygosity and paralogous sequence

ExcessHet is the phred-scaled P value of an exact test for excess heterozygosity computed by GATK at joint genotyping [3]. The GATK joint-genotyping workflow filters sites with ExcessHet above 54.69 ($P = 3.4 \times 10^{-6}$) [4]. The supplied VCF carries the annotation but not the filter. Sites above 54.69 were excluded from all primary analyses (Table S11). Of the 1,454 baseline higher-frequency contrasts, 564 were ExcessHet-flagged and 359 were removed by the read-support mask (Figure S4).

The VWF and CNN2 alleles annotated as pathogenic lie in runs of adjacent sites with ExcessHet 160, no homozygous-alternate calls and median allele fractions of 0.13 to 0.29. The VWF sites lie in exon 28, within the region duplicated in VWFP1 [5, 6] (Figure S6; Tables S10 and S10b).

#### Chromosome X, sex inference and relatedness

Non-pseudoautosomal X was GRCh38 X excluding 10,001–2,781,479 and 155,701,383–156,030,895. Sex was inferred from three measures: X heterozygosity (threshold 0.0383), the X-to-autosome depth ratio and median depth at male-specific Y sites (Figures S7–S9). Recorded and inferred sex disagreed for three participants (genetic sex 108 female, 91 male). X-chromosome ploidy was assigned from genetic sex, and male genotypes contributed one allele. For G6PD p.Ser188Phe in the 187 unrelated participants, this gave 4 of 289 X chromosomes (nominal $P = 0.013$ against MID 183/4,353); using recorded sex gave 4 of 288 ($P = 0.013$), and excluding the three sex-discordant participants gave 4 of 284 ($P = 0.018$). The four carriers (two men, two women) had concordant sex.

Kinship was estimated as KING-robust kinship [7, 8] on 32,622 LD-pruned autosomal SNVs ($r^2 < 0.2$, 500-kb windows). Fifteen pairs exceeded 0.0442: 8 first-degree, 1 second-degree and 6 third-degree. One member of each pair was removed greedily, leaving 187 unrelated participants. An earlier estimate on 35,002 markers with different pruning identified 12 pairs (188 unrelated); the 187 set is used throughout.

#### Principal-component analysis

The joint PCA used PLINK v2.0.0-a.6.9LM [7] on the cohort and 3,280 HGDP and 1000 Genomes samples [9] (EAS 729, AFR 680, CSA 673, EUR 666, AMR 378, MID 154). Of 5,552 shared markers, 3,176 remained after exclusion of long-range LD regions, a reference-only HWE filter, missingness and minor-allele-frequency filters, and pruning (–indep-pairwise 200 50 0.1). Ten components were computed. Eigenvalues for PC1 to PC4 were 111.6, 99.4, 33.0 and 28.5, that is 35.7%, 31.8%, 10.6% and 9.1% of the sum of the first ten. Each participant was assigned to the nearer of the Middle Eastern and European centroids in the first k components (Table S8, Figure S2).

#### Reference comparison and calibration of absence

Reference counts came from gnomAD v4.1 joint exome and genome data, matched by exact GRCh38 chromosome, position, reference and alternate allele [10]. Each filtered cohort allele was assigned to one of four categories (Table S20): - observed in MID (MID AC >0); - callable absence (MID AN >0 and AC = 0); - zero MID allele number; - no MID record.

The last category includes loci without a gnomAD record. gnomAD all-sites allele-number files were not queried, so this category does not establish that a locus was uncovered.

To calibrate callable absence, only autosomal sites were used (9,950 filtered alleles on chromosome X were excluded), and the 187 unrelated participants were split at random into a discovery half of 94 and a replicate half of 93, ten times. Each person contributes two allele copies at an autosomal site, so the replicate half contains 186 allele copies. gnomAD reports allele counts and allele numbers but not individual genotypes. For each allele seen in the discovery half, we therefore computed the probability that none of 186 allele copies drawn at random, without replacement, from MID would carry it. This hypergeometric probability, C(AN − AC, 186)/C(AN, 186) from the MID allele count AC and allele number AN, corresponds to sampling 93 MID individuals. It was compared with the observed absence in the replicate half, by allele count in the discovery half and by variant type, at three site sets: all retained sites; sites callable (depth ≥10) in at least 189 of the original 199 participants, that is with at most ten missing genotypes in the whole callset; and, as a stricter missingness sensitivity, sites callable in every member of the 93-person comparison half, so that absence there cannot arise from a missing genotype (Table S21; Table S21c; Figure 2B; Figure S14). Splits were drawn with NumPy default_rng(seed 11); the earlier parametric calibration used seed 7. Table S21c gives the mean, SD and range of each ratio across the ten splits. Because the splits reuse the same 187 people, this range describes split-to-split variability and is not a sampling interval. For SNVs overall, the ratio of MID to cohort absence was 0.98 (range 0.97–0.99) at all sites, 1.02 (1.01–1.03) at sites callable in ≥189 participants and 1.02 (1.01–1.03) at fully callable sites; for alleles seen one to five times at fully callable sites it was 1.01 to 1.12 for SNVs and 0.98 to 1.17 for indels; for alleles seen at least six times it was 2.3 (1.8–2.7) for SNVs. Among the 187 unrelated participants, 13.8% of autosomal alleles with a MID allele number were absent from MID (Jeffreys expectation 1.5%), and 360 alleles seen at least six times (284 SNVs, 76 indels) were absent, none ClinVar P/LP. In a sensitivity analysis that also included chromosome X (counting two allele copies per man, which overstates male X copies), ratios for alleles seen one to five times differed by less than 0.01; the stratum of alleles seen at least six times, which rests on few absent alleles, changed more (for example, indels at well-called sites 3.4 versus 3.6). This calibration compares absence at equal sample size; it does not estimate what fraction of absence from the full MID group is due to sampling, and similar average rates do not imply similarity at each locus.

For comparison, beta-binomial expectations were computed with a Jeffreys prior: $\Pr(\text{ACref} = 0 \mid k, n, m) = B(k + 0.5, n - k + 0.5 + m)/B(k + 0.5, n - k + 0.5)$. Here $k$ of $n$ cohort alleles carry the variant and $m$ is the reference allele number. A beta prior fitted by maximum likelihood to the split data collapsed towards a point mass near zero frequency (shape parameter below $10^{-10}$). The beta family therefore does not describe the allele-frequency spectrum of cohort-discovered alleles.

#### Autozygosity

For ROH calling we selected autosomal biallelic SNVs with QUAL ≥30, ExcessHet ≤54.69, mean site depth ≥20, minor allele frequency ≥0.05 and fewer than 5% missing genotypes after setting genotypes with depth below 10 to missing (96,978 SNVs). ROH were called with bcftools roh using cohort allele frequencies. Sensitivity analyses split runs at marker gaps longer than 1 Mb, used alternative marker sets, counted long ROH only and computed F against gnomAD Middle Eastern allele frequencies (Table S13).

#### Frequency tests

Two-sided Fisher exact tests compared cohort allele counts with MID counts for the 93,724 coding or splice variants with baseline cohort AC ≥2 and MID AN >100. The Bonferroni threshold was 0.05/93,724 ($5.3 \times 10^{-7}$). The primary analysis used masked genotypes in the 187 unrelated participants with callable allele numbers, after excluding ExcessHet-flagged sites (Table S2b). At the 1,664 tested non-pseudoautosomal X sites, women contributed two allele copies and men one, with sex from genetic inference (102 women and 85 men among the 187), and heterozygous calls in men were treated as missing; counting male X genotypes as diploid, as in the supplied summary counts used for Table S2, would have given 487 rather than 474 higher-frequency contrasts (14 X-linked contrasts lost and 1 gained with sex-aware counting) and the same number of lower-frequency contrasts (287; 1 lost and 1 gained). Analyses in all 199 participants and with baseline genotypes are in Tables S1, S2 and S9 and Figures S5 and S11. The paralogous gene-family and segmental-duplication flags are defined in Table S2.

#### Annotation and cancer-gene classification

Annotation used OpenCRAVAT 3.1.1 [11] with the ClinVar release of 1 July 2026 [12]. In a 621-record check against an independent annotation, 578/578 protein HGVS and 324/328 ClinVar significance calls agreed.

Variants in 75 cancer-predisposition genes were assigned to categories using ClinGen gene–disease validity curations downloaded on 28 September 2026 [13] (Table S19): - established dominant predisposition, meaning a Definitive or Strong autosomal-dominant cancer curation (PALB2, PMS2, SDHB, ATM); - dominant association limited or refuted (FANCM, breast cancer, Limited; RAD50, breast and ovarian cancer, Refuted); - heterozygous carrier of a recessive syndrome (FANCC, MUTYH, NTHL1); - predicted loss of function without a ClinVar P/LP classification.

Table S19 also gives MID allele counts and numbers for each variant.

#### Clonal hematopoiesis tiering

Candidates were called in a separate somatic branch (depth ≥10, ≥3 alternate reads, no allele-fraction floor). The background was 223,253 singleton heterozygous genotypes from the same callset. For each candidate, background genotypes of the same variant type with depth within ±20% of the candidate's depth were selected. The empirical percentile of the candidate's allele fraction among them was recorded, together with a one-sided binomial P for allele fraction below 0.5 (Table S14b).

Tiers were defined as follows: - probable: binomial P <0.01 and allele fraction at or below the 2.5th matched percentile; - possible: binomial P <0.01 only; - candidate only: neither criterion met.

Table S14c gives, by depth and variant type, the fraction of background singletons flagged by the binomial criterion alone. For artefact checks, alternate reads at each candidate position were counted in the other 198 participants, and FS, SOR and MQ were recorded (Table S14).

#### Recessive genotypes, severity and carrier-screening threshold

Qualifying alleles were filtered ClinVar P/LP alleles with at least two review stars in the 1,172 green biallelic autosomal genes of PanelApp Australia panel 3139 v1.0 (Mackenzie's Mission) [14]. Each allele was mapped to its ClinVar condition and flagged if it is hypomorphic or of variable severity (for example mild hyperphenylalaninemia PAH alleles, CFTR 5T and SERPINA1 deficiency alleles), or if it causes disease in heterozygotes (for example LDLR, KCNQ1 and RYR1) (Table S12e). The flags are provisional pending clinical-genetic review.

For gene $g$ with $K\text{g}$ qualifying allele copies among $M$ = 374 chromosomes, the pairwise probability was $K\text{g}(K\text{g} - 1)/[M(M - 1)]$. This estimates $p\text{g}^2$ without pairing a chromosome with itself and is unbiased if the 374 chromosomes are treated as a simple random sample. Participants, not chromosomes, were sampled. When each carrier has one qualifying allele in the gene and nobody is homozygous, pairing distinct people gives $C\text{g}(C\text{g} - 1)/[4N(N - 1)]$ for $C\text{g}$ carriers among $N$ = 187, which is larger by $(2N - 1)/[2(N - 1)]$ = 373/372 (baseline 2.0 per 1,000 either way). These conditions hold here: no participant was homozygous and none carried two qualifying alleles in the same gene (155 allele copies in 155 carrier–gene pairs), so phase did not arise. Missing genotypes were counted as non-carriers. The primary unrelated analysis contains 129 qualifying alleles (those carried by at least one of the 187 unrelated participants). Of these, 125 were callable in at least 178 of the 187 (113 in all 187; median 187), and four (GAA c.-32-13T>G, FKTN p.Arg307Ter, DOLK p.Met1?, FTCD p.Pro331AlafsTer2) in 93 to 165. The recessive rows of Table S21b list 134 alleles because they cover all 199 participants; the five additional alleles (in KCNQ1, ITGB4, ALPL, COL4A3 and FANCL) are carried only by related participants and are flagged with in_primary_unrelated_set = False. The modeled proportion of offspring with a qualifying biallelic genotype at inbreeding coefficient $F$ was $\Sigma\text{g}[(1 - F)K\text{g}(K\text{g} - 1)/\{M(M - 1)\} + F\,K\text{g}/M]$. Summing over genes approximates the probability of at least one qualifying biallelic genotype when per-gene probabilities are small and genes are independent. The outputs assume random union of the observed chromosomes and full penetrance of qualifying genotypes, omit alleles not seen in 187 people, and depend on the gene list, ClinVar release and severity flags; they are conditional model outputs, not estimates of affected births. At-risk couples were the proportion of the 17,391 pairs of distinct unrelated participants who carry qualifying alleles in the same gene.

Delete-one jackknife standard errors gave 95% intervals, on the log scale for ratios and small proportions (Table S12f). The participant bootstrap was not used, because resampling duplicates carriers and inflates pairwise estimates.

Gene carrier frequencies (carriers/187), with Wilson intervals, were compared with the ACMG threshold of 1/200 [15], once with all qualifying alleles and once with only the alleles not flagged for variable severity. MID values for the same cohort-observed alleles were computed as $2p(1 - p)$ from their summed MID allele frequency (Table S12g). They are not gene-level MID carrier frequencies, which would include pathogenic alleles not observed here, and they cannot show that a gene falls below 1/200 in MID; gene-level MID frequencies were not computed. Lower bounds are fragile at these counts: 3 of 187 carriers gives a Wilson lower bound of 0.55%, 2 of 187 would give 0.29%.

#### Secondary findings

Candidates were P/LP variants in the 84 genes of ACMG SF v3.3 [16]. For each candidate, ClinVar was re-queried on 28 September 2026 for classification, review status and asserted conditions (Table S16). The following were not counted: - single heterozygous variants in GAA and MUTYH; - heterozygous ABCD1 variants in women; - variants with zero review stars; - probable or possible clonal variants, and any finding whose somatic or constitutional origin is unresolved (the TP53 frameshift), which is withheld from the germline estimate; - variants whose asserted condition did not match the ACMG SF condition (for example, RYR1 variants not asserted for malignant hyperthermia susceptibility).

The primary estimate required at least two review stars and an exact match between the asserted ClinVar condition and the ACMG SF condition. KCNQ1 c.921G>A (p.Val307=; VCV 53127) is asserted for Jervell and Lange-Nielsen syndrome and cardiovascular phenotype, not explicitly for long QT syndrome type 1; it is reported as unresolved and excluded from the primary estimate of 8 participants (4.0%), giving 9 (4.5%) if included. TTN was restricted to truncating variants. The one TTN candidate (GRCh38 chr2:178532361 G>A; NM_001267550.2 c.104254C>T, p.Gln34752Ter; stop-gained; ClinVar VCV004085005, likely pathogenic, one star, condition not provided) does not meet the exact condition-match rule and its cardiac-exon usage was not assessed; it is reported as unresolved. With at least one star, 9 participants meet the rule (the primary 8 plus ENG p.Pro555LeufsTer14, asserted for ENG-related disorder); including the TTN candidate, who carries no other finding, gives 10. No finding has been confirmed orthogonally.

#### MEFV definitions and FROH cross-tabulations

The primary MEFV carrier definition used six classic exon 10 alleles (V726A, M680I, M694V, M694I, R653H, A744S). A strict definition used four (V726A, M680I, M694V, M694I), and E148Q was reported separately (Table S13d). Table S13c cross-tabulates

FROH classes (<1/64, 1/64 to <1/16, ≥1/16) with each finding category and gives ROH counts and lengths by class.

#### Frequency screen

The screen used the 20,377 protein-altering records annotated as of uncertain significance or with conflicting interpretations (Table S6a). The proxy maximum was 0.001 for autosomal-dominant or X-linked inheritance strings and 0.01 otherwise. A record was retained when the one-sided 95% lower bound of its cohort frequency (5th percentile of Beta(AC, AN − AC + 1)) exceeded the proxy and the MID frequency did not (Tables S6 and S6b; Figure S12).

#### Data, code and reproducibility

Figure source data contain aggregate values or anonymized per-participant metrics without identifiers. Genotype-level reproduction requires the controlled VCF. The analysis and figure code accompanies the submission (Python 3.12; bcftools 1.24; PLINK v2.0.0-a.6.9LM; cyvcf2 0.34.0; NumPy 2.5.3; pandas 3.0.6; SciPy 1.18.1; statsmodels 0.15.0).

### Figure S1. Per-sample quality metrics by recruitment center

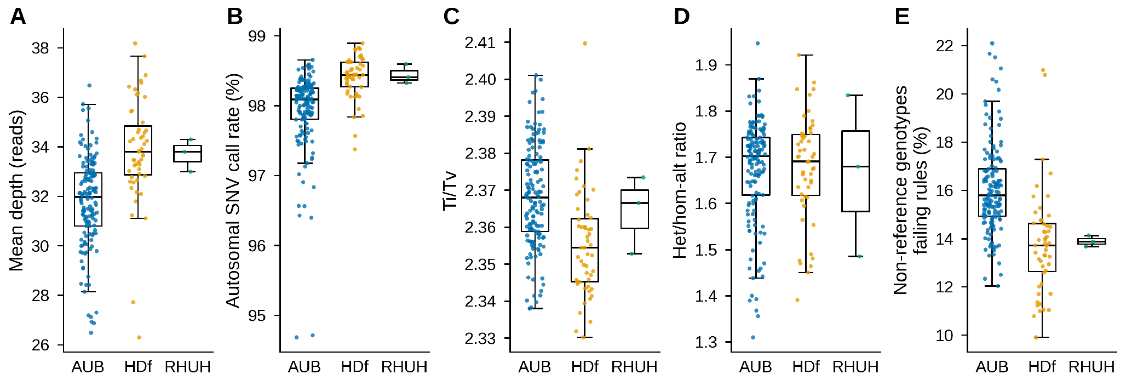


A, mean genotype depth. B, autosomal SNV call rate. C, transition/transversion ratio. D, heterozygous to homozygous-alternate ratio. E, percentage of non-reference genotypes failing any read-support rule. Boxes show median and interquartile range with whiskers to 1.5 times the interquartile range; points are participants. AUB, American University of Beirut Medical Center; HDf, Hôtel-Dieu de France Hospital; RHUH, Rafik Hariri University Hospital.

### Figure S2. Sensitivity of ancestry summaries

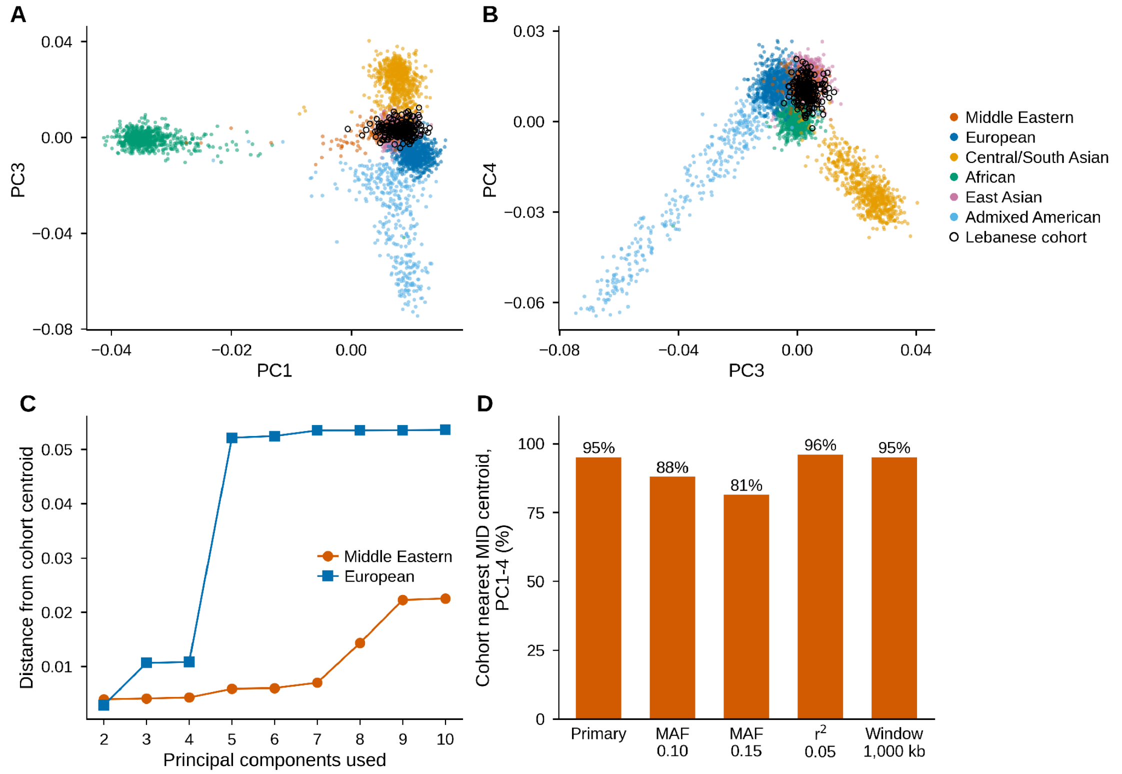


A, PC1 against PC3 and B, PC3 against PC4 from the joint principal-component analysis; cohort participants are open circles. C, distance from the cohort centroid to the Middle Eastern and European reference centroids by the number of components used. D, percentage of participants nearest the Middle Eastern centroid in PC1 to PC4 for marker sets defined by alternative minor-allele-frequency, LD $r^2$ and window thresholds.

### Figure S3. Genotype read-support audit

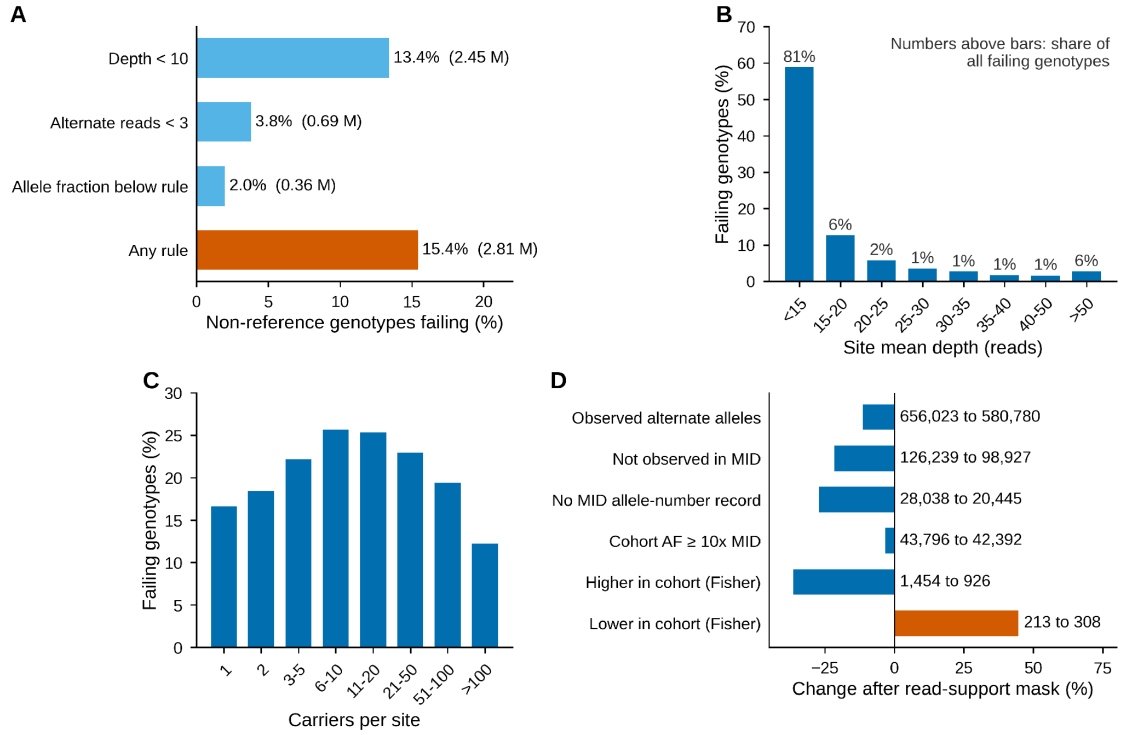


A, percentage of the 18.2 million non-reference genotypes failing each read-support rule and any rule. B, failing genotypes by site mean depth; numbers above bars give each bin’s share of all failing genotypes. C, failing genotypes by number of carriers per site. D, change in allele and contrast counts after applying the read-support mask in all 199 participants, before ExcessHet filtering.

### Figure S4. Fate of frequency contrasts under quality filters

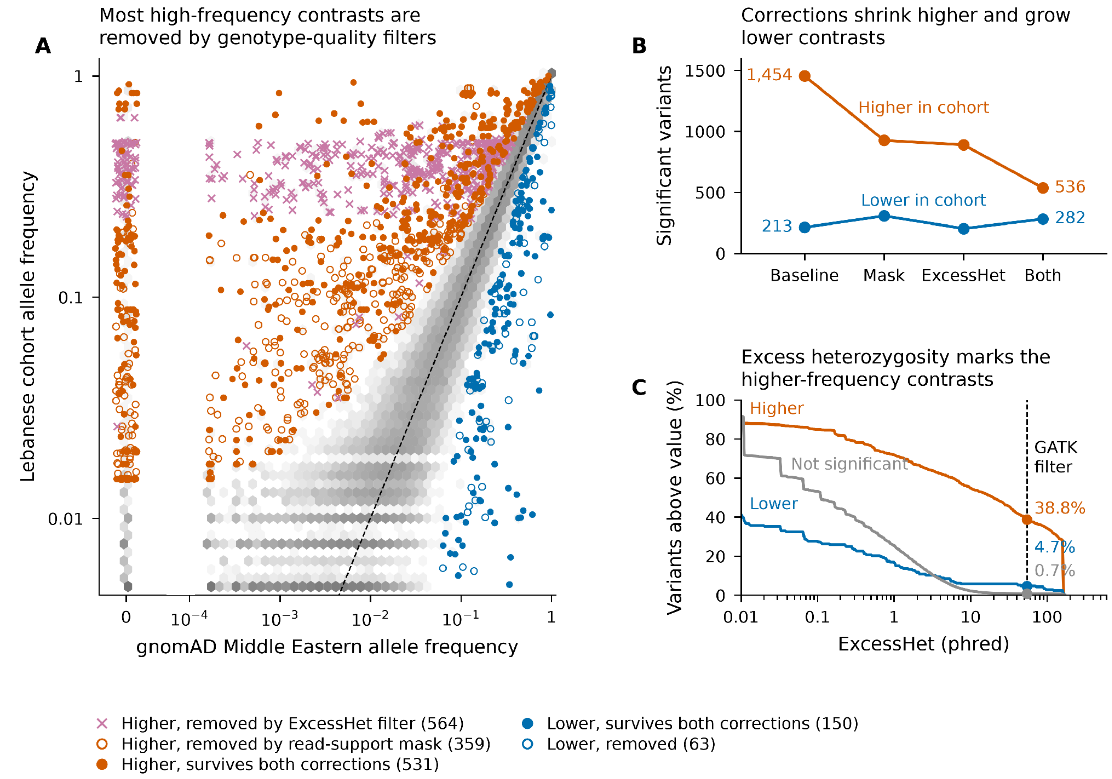


A, cohort (all 199 participants) against gnomAD Middle Eastern allele frequency for the 93,724 tested variants. Baseline higher-frequency contrasts are marked by whether they were removed by the ExcessHet filter (crosses), removed by the read-support mask (open circles) or survived both corrections (filled circles); lower-frequency contrasts are blue. B, significant contrasts at baseline, after the mask, after the ExcessHet filter and after both. C, cumulative distribution of ExcessHet among higher, lower and non-significant variants; the dashed line marks the GATK threshold of 54.69. The primary analysis in 187 unrelated participants is in Figure 2A and Table S2b.

### Figure S5. Sensitivity of frequency contrasts

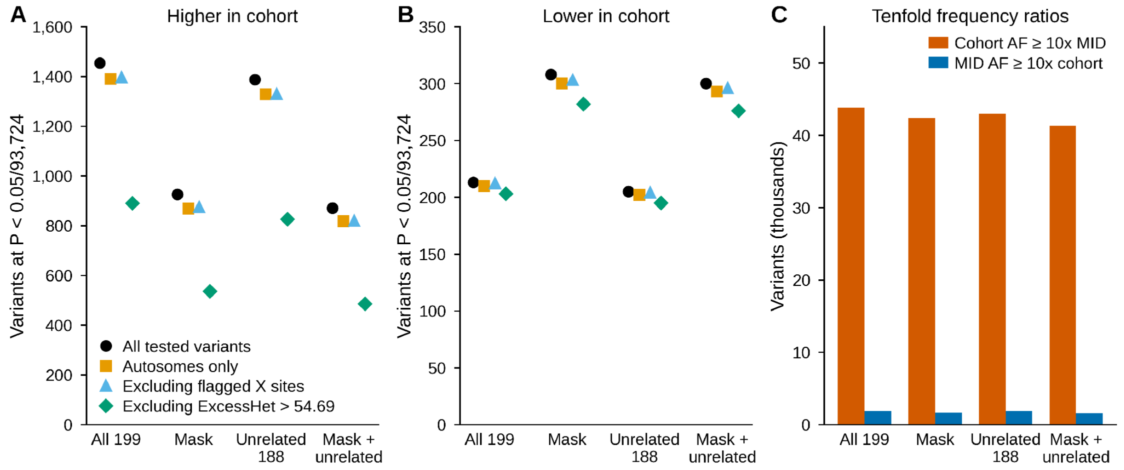


A, variants with higher and B, lower cohort frequency than the gnomAD Middle Eastern group at P <0.05/93,724 for all tested variants, autosomes only, after excluding X sites with male heterozygous calls, and after excluding sites with ExcessHet >54.69. Each is shown in four analyses: all 199, masked, the 188 unrelated participants of the first kinship estimate, and masked plus unrelated (this sensitivity figure uses the 188 unrelated participants of an earlier kinship estimate on 35,002 markers; the primary analysis in 187 unrelated participants is in Figure 2A and Table S2b). C, variants with at least tenfold higher cohort or Middle Eastern allele frequency.

### Figure S6. Paralog artefacts at VWF and CNN2

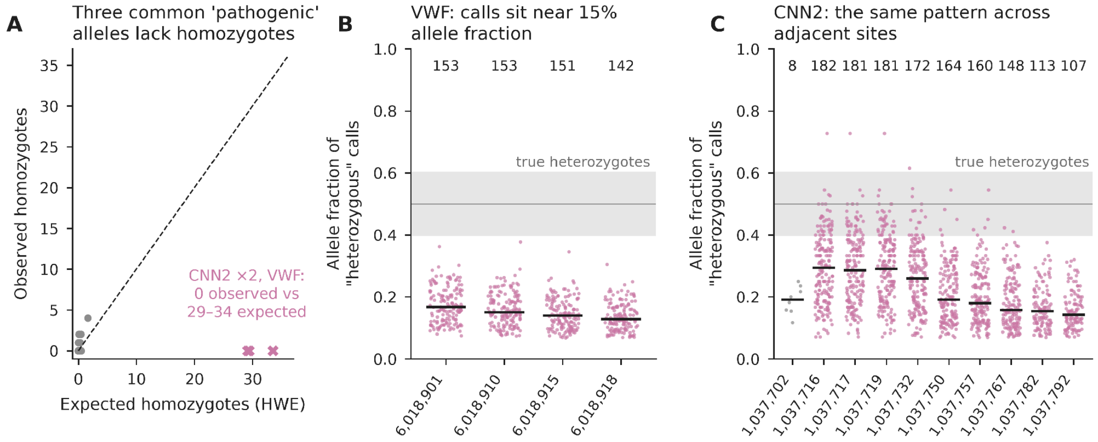


A, observed against expected homozygotes under Hardy–Weinberg proportions for the recurrent pathogenic-annotated alleles; CNN2 p.Gly263Ser, CNN2 p.Arg266Gln and VWF p.Ser1506Leu (crosses) have no homozygote where 29 to 34 were expected. B, alternate-allele fraction of heterozygous calls at four adjacent VWF exon 28 sites and C, at ten adjacent CNN2 sites; numbers give heterozygous calls per site, bars mark medians, and the shaded band marks the range expected for true heterozygotes.

### Figure S7. Sex-chromosome inference

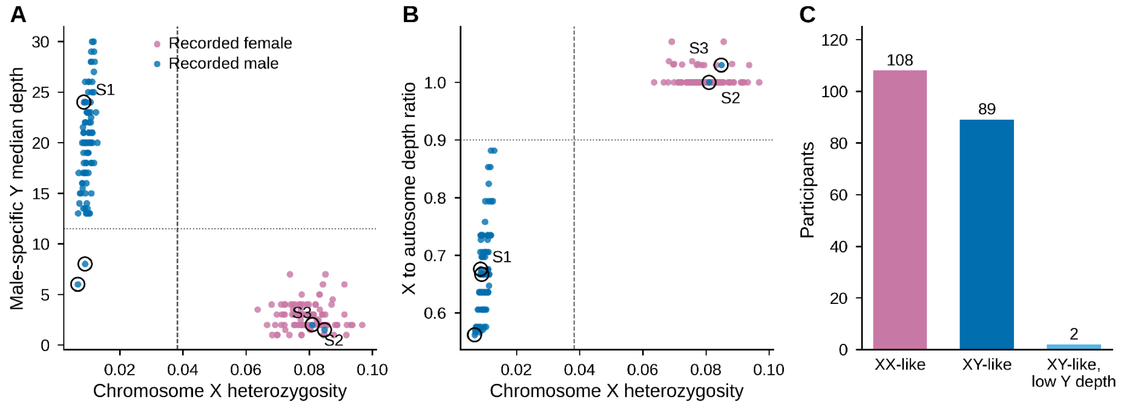


A, male-specific Y median depth and B, X-to-autosome depth ratio against non-pseudoautosomal X heterozygosity; colors give recorded sex, and dashed and dotted lines mark the inference thresholds. Circled points are the three discordant participants (S1 to S3) and two XY-like participants with low Y depth. C, participants by inferred sex-chromosome class.

### Figure S8. Heterozygous calls in males on non-pseudoautosomal chromosome X

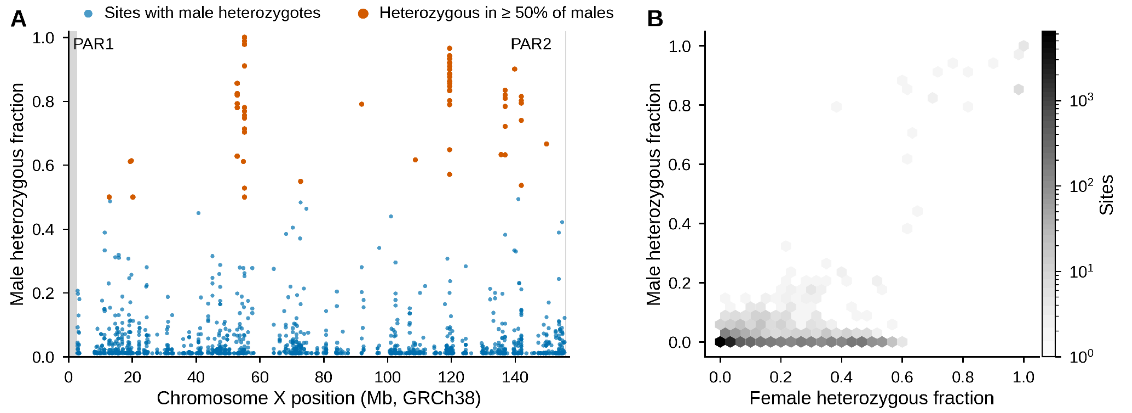


A, fraction of inferred males heterozygous at each non-pseudoautosomal X site along the chromosome; sites at which at least half of males are heterozygous are vermilion, and pseudoautosomal regions are shaded. B, fraction of females against fraction of males heterozygous at each site (hexagonal bins).

### Figure S9. Participants with discordant recorded and inferred sex

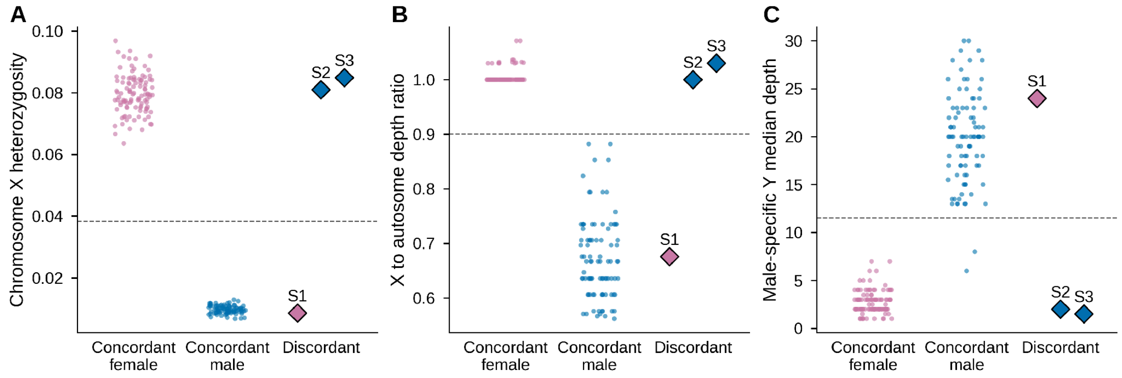


A, chromosome X heterozygosity, B, X-to-autosome depth ratio and C, male-specific Y median depth for concordant females, concordant males and the three discordant participants (S1, recorded female; S2 and S3, recorded male). Identifiers are replaced by S1 to S3.

### Figure S10. Pooled gene-level allele-frequency ratios

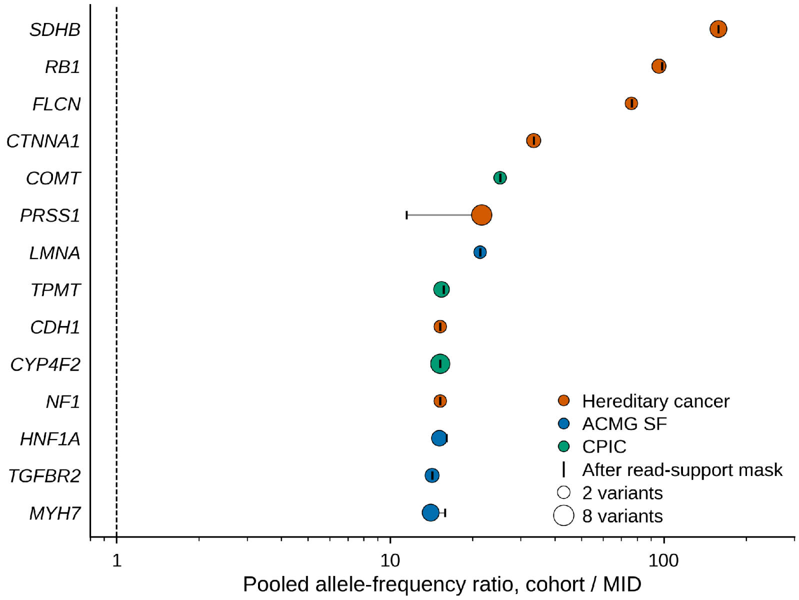


Pseudocount ratio of pooled cohort to pooled Middle Eastern allele frequency for 14 panel genes (log scale), computed over rare protein-altering variants as $[(\Sigma AC + 0.5)/(\Sigma AN + 1)]$ for the cohort divided by the same quantity for the Middle Eastern group. Symbol area scales with the number of variants, and color gives the gene panel; ticks mark the ratio after the read-support mask, and the dashed line marks a ratio of 1. For genes without Middle Eastern alternate alleles, ratios are determined largely by the pseudocount. Descriptive only.

### Figure S11. Reference status and frequency contrasts in unfiltered data

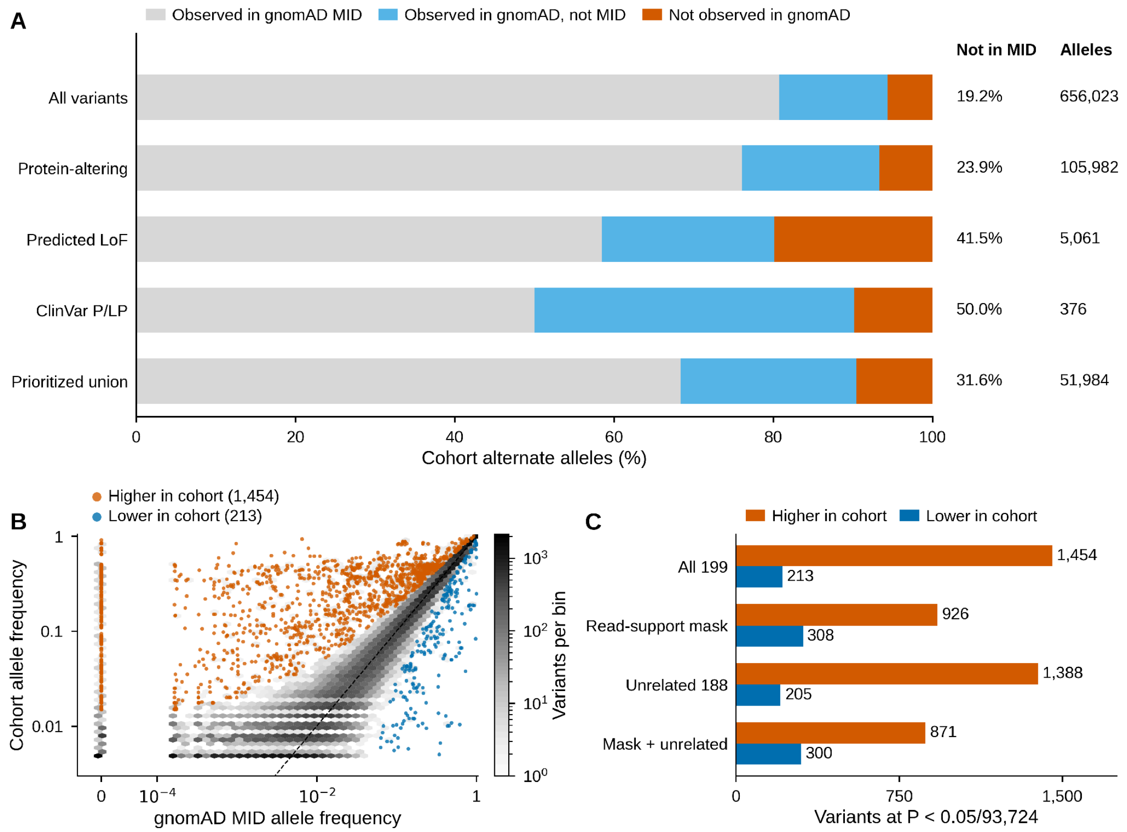


A, reference status of cohort alternate alleles in unfiltered data by annotation class. The proportion not observed in MID is not calibrated for sample size; the calibrated comparison is in Figure 2B and Figure S14. B, cohort against MID allele frequency for the 93,724 tested variants with baseline genotypes. C, significant contrasts in all 199 participants, after the read-support mask, in the 188 unrelated participants of the first kinship estimate, and with both. These unfiltered results are superseded by Figure 2A and Table S2b.

### Figure S12. Generic frequency screen

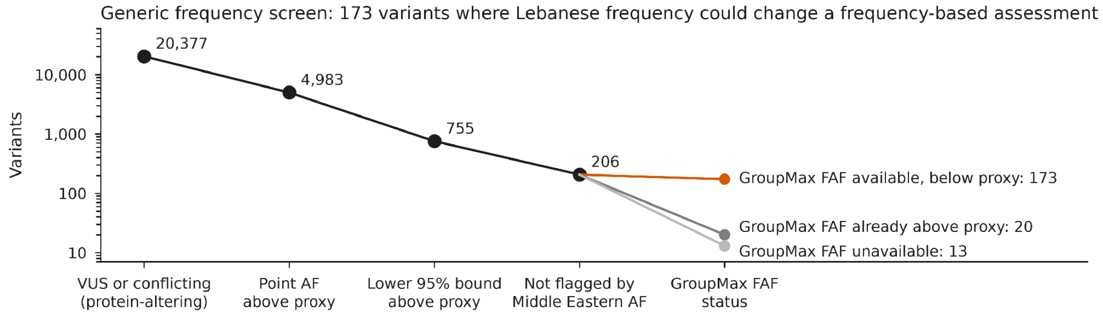


Number of protein-altering variants of uncertain significance or with conflicting classifications retained at each step of a screen comparing cohort frequency with a generic disease-frequency proxy (0.001 for dominant or X-linked, 0.01 otherwise). The steps are: point frequency above the proxy; one-sided 95% lower bound above the proxy; not flagged by the Middle Eastern frequency; and status of the GroupMax filtering allele frequency.

### Figure S13. Sample quality control of the supplied callset

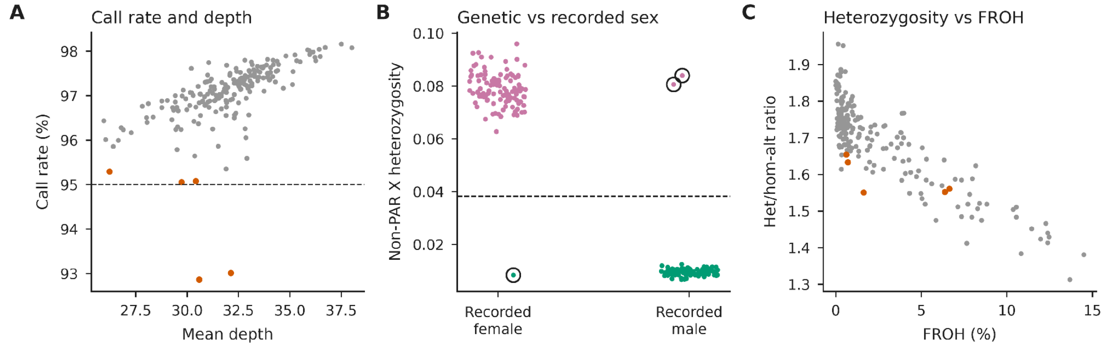


A, autosomal SNV call rate against mean depth; vermilion points are the five samples with the lowest call rates, and the dashed line marks 95%. B, non-pseudoautosomal X heterozygosity by recorded sex, colored by inferred sex (pink, female; green, male); circles mark the three discordant participants, and the dashed line marks the inference threshold. C, heterozygous to homozygous-alternate ratio against FROH; vermilion as in A. Source data: Table S17.

### Figure S14. Calibration of absence from the Middle Eastern reference

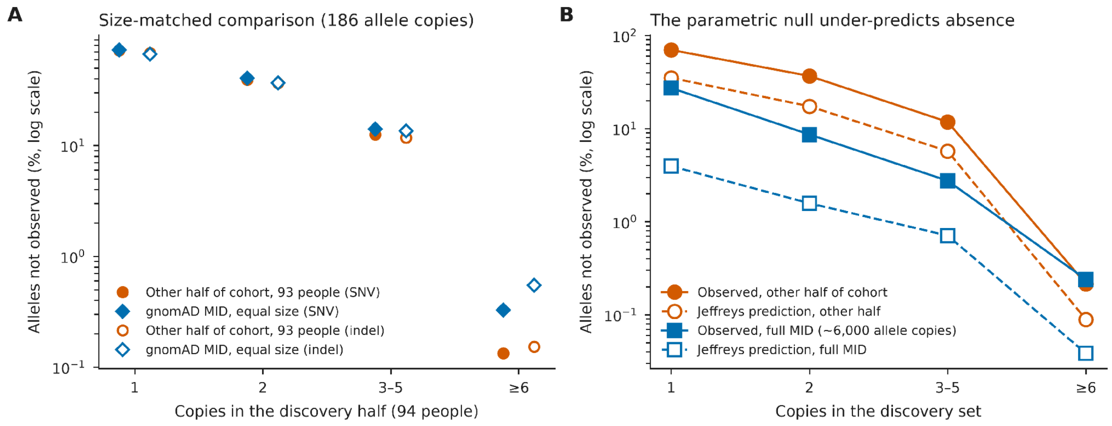


A, the 187 unrelated participants were split at random into halves of 94 and 93 people (ten splits). For alleles seen in the 94-person half, the percentage absent from the 186 allele copies of the other half (vermilion circles) is compared with the percentage expected to be absent from 186 allele copies drawn at random from gnomAD Middle Eastern (MID), the equivalent of 93 people (blue diamonds). Results are shown by number of copies in the discovery half; filled symbols are SNVs and open symbols indels; only autosomal sites callable (depth ≥10) in at least 189 of the original 199 participants are included; mean of ten splits (seed 11), with split-to-split ranges in Table S21c. B, observed absence (solid lines) and beta-binomial expectation with a Jeffreys prior (dashed lines), for the other half of the cohort (vermilion; discovery in 94 people) and for the full MID group (blue; about 6,000 allele copies; discovery in 187 people). Source data: Table S21.

#### Supplementary tables

Supplementary Tables are supplied as one Excel workbook (Supplementary_Tables_S1-S21.xlsx; one sheet per table) and, for large tables, as compressed CSV files.

| Table | Contents |
|---|---|
| S1 | Reference status, tenfold contrasts and frequency-test counts in four baseline analyses |
| S2 | Allelic Fisher tests for the 93,724 tested variants in all 199 participants, with frequency ratios and gene-family flags (supplied summary counts; male X genotypes counted as diploid, see S2b for sex-aware counts) |

| Table | Contents |
| --- | --- |
| S2b | Primary frequency tests in 187 unrelated participants with callable allele numbers (sex-aware X counts) |
| S3 | Descriptive gene-level frequency summaries |
| S4 | The 66 recurrent pathogenic-annotated alleles |
| S5 | Read support of secondary-finding candidates |
| S6, S6a, S6b | Frequency-screen candidates, inputs and threshold union |
| S7 | Reference status by annotation tier (unfiltered) |
| S8 | Nearest reference centroid by number of principal components |
| S9 | Frequency tests on autosomes only and after excluding X sites with male heterozygous calls |
| S10, S10b | Hardy–Weinberg exact tests for recurrent alleles in all and in unrelated participants |
| S11 | ExcessHet values and flags for the tested variants |
| S12 | Recessive carrier proportions under alternative definitions |
| S12b | Published benchmarks for recessive carrier frequency and consanguinity risk |
| S12c | External severe-recessive gene list analysis in all and in unrelated participants |
| S12d | Carrier genes under the external list, with MID frequencies |
| S12e | Variant–condition map with severity and heterozygote-phenotype flags |
| S12f | Recessive risk estimators with jackknife 95% intervals |
| S12g | Gene carrier frequencies against the ACMG 1/200 threshold; MID |

| Table | Contents |
|---|---|
| | values for cohort-observed alleles only (not gene-level) |
| S13, S13b | Autozygosity method sensitivity and Lebanese exome ROH thresholds |
| S13c | Findings and ROH lengths by FROH class |
| S13d | MEFV carrier definitions |
| S14, S14b, S14c | Clonal hematopoiesis artefact checks, candidate tiers and calibration of the binomial criterion |
| S15 | Regional disease alleles in unrelated participants |
| S16 | Condition-specific secondary-finding audit |
| S17 | Per-sample quality control (anonymous codes) |
| S18 | Headline results after excluding defined sample sets (two below 95% call rate, five lowest call rates, three sex-discordant, eight combined) |
| S19 | Gene–disease validity audit of cancer variants, with MID counts |
| S20 | Reference-absence categories |
| S21, S21b, S21c | Calibration of absence from MID; callable allele numbers for headline alleles (X-linked alleles count one copy per man); split-to-split variability and callability sensitivity (with per-split values) |

#### Supplementary references


1. Hendi NN, Chakhtoura M, Al-Sarraj Y, et al. The genetic architecture of vitamin D deficiency among an elderly Lebanese Middle Eastern population: an exome-wide association study. Nutrients. 2023;15:3216. doi:10.3390/nu15143216.

2. Rahme M, Sharara SL, Baddoura R, et al. Impact of calcium and two doses of vitamin D on bone metabolism in the elderly: a randomized controlled trial. J Bone Miner Res. 2017;32:1486–1495. doi:10.1002/jbmr.3122.

3. Broad Institute. GATK tool documentation: ExcessHet. https://gatk.broadinstitute.org/hc/en-us/articles/360037261311-ExcessHet (accessed 27 September 2026).

4. Broad Institute. WARP JointGenotyping pipeline overview (excess_het_threshold 54.69). https://broadinstitute.github.io/warp/docs/Pipelines/JointGenotyping_Pipeline/README (accessed 27 September 2026).

5. Mancuso DJ, Tuley EA, Westfield LA, et al. Human von Willebrand factor gene and pseudogene: structural analysis and differentiation by polymerase chain reaction. Biochemistry. 1991;30:253–269. doi:10.1021/bi00215a036.

6. Ye S, Eura Y, Matsumoto M, Kokame K. Genetic analysis using long-read sequencing to overcome the difficulties in VWF gene. Res Pract Thromb Haemost. 2025;9:102888. doi:10.1016/j.rpth.2025.102888.

7. Chang CC, Chow CC, Tellier LC, et al. Second-generation PLINK: rising to the challenge of larger and richer datasets. GigaScience. 2015;4:7. doi:10.1186/s13742-015-0047-8.

8. Manichaikul A, Mychaleckyj JC, Rich SS, et al. Robust relationship inference in genome-wide association studies. Bioinformatics. 2010;26:2867–2873. doi:10.1093/bioinformatics/btq559.

9. Koenig Z, Yohannes MT, Nkambule LL, et al. A harmonized public resource of deeply sequenced diverse human genomes. Genome Res. 2024;34:796–809. doi:10.1101/gr.278378.123.

10. gnomAD Production Team. gnomAD v4.1. Release announcement, 19 April 2024. https://gnomad.broadinstitute.org/news/2024-04-gnomad-v4-1 (accessed 15 September 2026).

11. Pagel KA, Kim R, Moad K, et al. Integrated informatics analysis of cancer-related variants. JCO Clin Cancer Inform. 2020;4:310–317. doi:10.1200/CCI.19.00132.

12. Landrum MJ, Lee JM, Benson M, et al. ClinVar: improving access to variant interpretations and supporting evidence. Nucleic Acids Res. 2018;46:D1062–D1067. doi:10.1093/nar/gkx1153.

13. Strande NT, Riggs ER, Buchanan AH, et al. Evaluating the clinical validity of gene-disease associations: an evidence-based framework developed by the Clinical Genome Resource. Am J Hum Genet. 2017;100:895–906. doi:10.1016/j.ajhg.2017.04.015.

14. Kirk EP, Ong R, Boggs K, et al. Gene selection for the Australian Reproductive Genetic Carrier Screening Project ('Mackenzie's Mission'). Eur J Hum Genet. 2021;29:79–87. doi:10.1038/s41431-020-0685-x.

15. Gregg AR, Aarabi M, Klugman S, et al. Screening for autosomal recessive and X-linked conditions during pregnancy and preconception: a practice resource of the

American College of Medical Genetics and Genomics (ACMG). Genet Med. 2021;23:1793–1806. doi:10.1038/s41436-021-01203-z.

16. Lee K, Abul-Husn NS, Amendola LM, et al. ACMG SF v3.3 list for reporting of secondary findings in clinical exome and genome sequencing: a policy statement of the American College of Medical Genetics and Genomics (ACMG). Genet Med. 2025;27:101454. doi:10.1016/j.gim.2025.101454.
